# Accounting for cross-registration in monitoring responsible research in clinical trials: A cross-sectional study of trials at German university medical centers

**DOI:** 10.1101/2025.04.01.25325043

**Authors:** Luciano Fernandez, Elisa Bascunan Atria, Samruddhi Yerunkar, Maia Salholz-Hillel, Vladislav Nachev, Susanne Gabriele Schorr, Daniel Strech, Delwen L. Franzen

**Author notes:** **Corresponding author:** Delwen L. Franzen. These authors contributed equally to this work (Luciano Fernandez, Elisa Bascunan Atria).

## Abstract

**Background:** Identifying and monitoring clinical trials is crucial for responsible research systems and is relevant to all stakeholders in clinical research. Trial registries support accountability and transparency by providing a publicly available and rich source of information on trials. Cross-registration, where the same trial is registered in more than one registry, can complicate efforts to study the clinical research enterprise. Duplicate entries may need to be de-duplicated or merged, and discrepancies across registrations may affect conclusions about trial adherence to registration and reporting standards.

**Methods:** We identified and characterized cross-registrations in an existing dataset of 2,895 trials conducted at German university medical centers and registered on ClinicalTrials.gov or the German Clinical Trials Register. We focused on cross-registrations on the EU Clinical Trials Register (EUCTR), given the legal basis of registration and reporting in this registry. We manually validated a sample of EUCTR cross-registrations, and assessed discrepancies in key information across registrations of the same trial.

**Results:** We identified 625 potential EUCTR cross-registrations. The majority of potential cross-registrations (75%, n = 470) were linked through a Trial Registration Number (TRN) in at least one of the registries. Only 17% (n = 109) were linked through a TRN in both registries. Title matching and TRNs in trial results publications led to an additional 155 (25%) potential cross-registrations. For manually validated EUCTR cross-registrations (n = 228), we found discrepancies in the registration status, recruitment status, completion date, and availability of summary results across registration of the same trial.

**Conclusion:** Unambiguous identification of trials and consistent trial records are essential to support the inclusion of relevant trials in systematic reviews, and for monitoring responsible research. Yet, registrations of the same trial are often inadequately linked and present discrepant information. Our findings suggest the need for additional guidance and coordination on cross-registration across stakeholders.

## Introduction

Clinical trials are the cornerstone of evidence-based medicine. Patients put themselves at risk to advance medical knowledge, and the results of clinical trials are used to inform medical decision-making. Despite their significance to human health, there is long-standing evidence that many trials fall short of best practices for transparency. For instance, studies have shown that trial results are often incomplete, delayed, or not reported at all, hampering evidence synthesis and distorting the understanding of the medical evidence base (1–5). Trial registries as well as international standards, laws, and policies for trial registration and reporting (6–9) serve as important mechanisms to support transparency, combat bias, and maximize the value of trials. Ensuring trials adhere to transparency standards, and responsible research practices more broadly, is relevant to all stakeholders in clinical research, including funders, research performing institutions, industry, (meta-)researchers, regulatory bodies, clinicians and patients.

Monitoring responsible research in clinical trials is an important step in supporting compliance and requires a comprehensive overview of studies and information about these studies. Clinical trial registries support accountability and transparency in medical research by providing a rich and publicly available source of information on planned, ongoing, and completed clinical studies. The World Health Organization’s (WHO) Registry Network includes 17 primary registries, which together with ClinicalTrials.gov, house registrations for trials conducted across different jurisdictions. The WHO International Clinical Trials Registry Platform (ICTRP) serves as a meta-registry and combines data from primary registries and data providers. Registries serve as a crucial resource not only to inform patients and the public of ongoing trials, but also to study the clinical research enterprise, monitor responsible research practices, and inform research improvement activities (10,11). However, the validity and impact of these activities depends on the quality of registry data (12) and how registries are being used as data sources in research activities (13).

A trial may be registered on more than one registry, which we here refer to as “cross-registration”. In some cases, cross-registration may be required to meet regulatory obligations. For instance, a medicinal product trial approved under the Clinical Trials Directive with enrollment in the European Union and seeking market approval in the United States must be registered on both ClinicalTrials.gov and the EU Clinical Trials Register (EUCTR). Furthermore, the trial is legally required to report results on both registries. Following the Clinical Trials Regulation 536/2014 (14), EU drug trials ongoing as of 30 January 2025 must be registered and report results on the Clinical Trial Information System (CTIS), the successor to EUCTR. In other cases, duplicate trial registration may occur due to a lack of coordination between organizations and investigators participating in multi-site trials (15), or to meet additional requirements from regulators, funders, or institutions. Some trial registries also import trials from other registries with recruitment sites in a particular jurisdiction; for example, the Australian New Zealand Clinical Trials Registry imports trials registered on ClinicalTrials.gov with recruitment sites in Australia and New Zealand (16).

Cross-registration, as well as duplicate registrations within registries, can complicate efforts to study the clinical research enterprise. The number of registrations is greater than the number of trials, and poorly linked cross-registrations of the same trial can pose a challenge for determining how many trials actually exist (15). Depending on the research question, duplicate entries may need to be identified and subsequently deduplicated or merged, and variables may need to be harmonized across registries.

Discrepancies across registrations may also affect conclusions about trial adherence to registration and reporting standards: for example, a trial may have reported summary results in one registry but not in another. Previous work on registration trends in Germany found the same trials in more than one database, notably the German Clinical Trials Register (DRKS), ClinicalTrials.gov, and EUCTR (17). Thus, the trial landscape in Germany is particularly well-suited to gain a deeper understanding of cross-registrations.

In this study, we sought to identify and characterize cross-registrations in an existing dataset of trials conducted at German University Medical Centers (UMCs). Trials in this dataset are registered on ClinicalTrials.gov or the DRKS and are not limited to a specific intervention type. We focused on exploring cross-registration in the EUCTR, given the legal basis of registration and reporting in this registry (14). In a manually validated sample of these cross-registrations, we examined discrepancies in the registration status, recruitment status, completion date, and availability of summary results across registrations of the same trial. Our findings may inform future efforts to synthesize clinical evidence and monitor trials’ adherence to responsible research practices.

## Methods

This cross-sectional study followed a protocol (https://osf.io/yzxf3, 12 September 2024) and is reported according to the Strengthening the Reporting of Observational Studies in Epidemiology (STROBE) guideline for cross-sectional studies (18).

### Data collection

We used the following data sources to explore cross-registrations:

1. IntoValue, a dataset of 2,895 clinical trials conducted at German UMCs and related results publications (19). Trials in this dataset are registered on ClinicalTrials.gov (n = 2,253) or DRKS (n = 642) with a completion date between 2009 – 2017 per the registry. Trials include all interventional studies regardless of intervention type. We built on an initial set of potential EUCTR cross-registrations in IntoValue identified through Trial Registration Numbers (TRNs) in ClinicalTrials.gov and DRKS (registry download date: 1 November 2022) and in related publications. To assess prospective registration of IntoValue trials, we obtained the relevant registry fields from the Aggregated Analysis of ClinicalTrials.gov (AACT) and DRKS on 27 September 2024 and 20 December 2024, respectively.
2. Full EUCTR dataset, including protocols for all member states associated with a trial (download date: 3 February 2024) and results where available (download date: 15 February 2024).

We focused on cross-registrations between IntoValue trials (registered on ClinicalTrials.gov or DRKS) and EUCTR. Duplicate registrations between ClinicalTrials.gov and DRKS, or within the same registry, were outside the scope of this study. We required one of the following conditions for trials to be considered as potentially cross-registered: 1) both registrations listed a TRN pointing to the other registration, 2) one of the registrations listed a TRN pointing to the other registration, 3) the title of the trial was an approximate match on both registries, 4) a EUCTR identifier was mentioned in a results publication associated with a trial registered on ClinicalTrials.gov or DRKS, or 5) the same sponsor protocol code was listed on both registries. The approach to identify potential cross-registrations is detailed in **Supplement S1**. The associated code is openly available on GitHub: https://github.com/maia-sh/intovalue-data.

Additionally, we sought to compare our findings to a query of the Clinical Research Metadata Repository (MDR) of the European Clinical Research Infrastructure Network (ECRIN) (https://newmdr.ecrin.org/). The MDR is a tool that aggregates data across various sources to increase the discoverability of clinical studies and related data objects. We queried the MDR via the Application Programming Interface (API) for identifiers associated with trials in the IntoValue dataset (query date: 25 October 2024). The resulting identifiers were filtered for EUCTR TRNs. The associated code is openly available on GitHub: https://github.com/quest-bih/intovalue-crossreg.

### Inclusion and exclusion criteria

Prior to registering our protocol, we performed a pilot manual review of a random sample of potential EUCTR cross-registrations (n = 100). Based on this pilot manual review, we decided to exclude potential cross-registrations matching exclusively on sponsor protocol code (n = 296) from further manual validation due to low precision in the pilot (10%, n = 20) and feasibility constraints. In the pilot manual review, we encountered several potential cross-registrations involving a TRN that did not resolve on the registry and could therefore not be validated; these cases were removed and replaced with a new potential cross-registration to validate. We used several approaches to identify additional non-resolving registrations, which were excluded from further manual validation (see detailed information in **Supplement S1**). For the comparison with the MDR, we excluded all potential cross-registrations identified exclusively via a matching sponsor protocol code but included potential cross-registrations involving a non-resolving TRN, since we also expected these to be listed in the MDR.

### Manual review of cross-registrations

We manually screened a sample of potential EUCTR cross-registrations. The sample was not randomly drawn across the entire dataset, but was informed by the characteristics indicating a cross-registration (e.g., title matching). We grouped potential cross-registrations into hierarchical categories based on these characteristics, and used sample size calculations and a prespecified stopping rule informed by the pilot manual review to determine the number of cases to manually review in each category (see protocol: https://osf.io/yzxf3). Cross-registrations linked exclusively via a TRN in a publication were all manually screened due to lower precision. A single coder (EBA) determined whether registrations were cross-registered, i.e., related to the same trial, based on matching on the following registry characteristics: title or sponsor, outcome (primary), study design (intervention model, masking, randomization), intervention, comparator (if applicable), planned patient enrolment, and indication/population. Whether a trial was cross-registered was left to the discretion of the coder (EBA); not all trial characteristics had to be an exact match. Edge cases were discussed with a second coder (SY, SGS) and discussed in the team as needed.

During the manual review, the coder (EBA) extracted the following trial characteristics across registrations: completion date, recruitment status, and the availability of summary results in the registry. These trial characteristics are relevant for identifying trials and for transparency audits; furthermore, comparing these characteristics across registrations of a trial provides insight into the quality of registry data. Extraction followed the aforementioned pre-registered protocol. For EUCTR, global completion dates from results sections were preferred for being the most recently updated. To facilitate comparison across registries, all completion dates were rounded to year-month. We mapped recruitment statuses across registries to one of the following overall recruitment statuses: “Ongoing”, “Completed”, or “Other” (**Supplement S2**). As a deviation to the protocol, only structured tabular results posted in the format of the host registry were considered as “results posted” in the main analysis. This decision was taken to highlight structured results reports; however, we note that DRKS did not provide a structured tabular results format directly on the registry at the time of the study, and even if registries provide this format, results are sometimes uploaded in other forms. In the case of summary results in a non-structured format (e.g., synopsis, linked publication), we checked the type of report but did not perform a manual review to confirm these reported on the results of the trial (**Supplement S2)**.

### Analysis

We report descriptive statistics for the number of potential EUCTR cross-registrations in the IntoValue dataset, and how they were identified (e.g., title matching). We also report the overlap of potential EUCTR cross-registrations identified through our methods and based on the ECRIN MDR. For a subset of potential cross-registrations that were manually screened during the pilot and main phases of data collection, we report the proportion of confirmed cross-registrations. For cross-registrations that were confirmed in the manual screening, we report the registration status, recruitment status, completion date, and availability of summary results across registrations of the same trial. The analysis of recruitment status was limited to confirmed cross-registrations with an available recruitment status in both registries. The analysis of completion date was limited to confirmed cross-registrations considered as “Completed” (includes completed and terminated trials, see **Supplement S2**) with an available completion date in both registrations. EUCTR trials without a protocol for Germany were excluded from the analysis of recruitment status and completion date (unless a completion date was provided in the results), as these variables may differ across country protocols. The analysis of summary results reporting was limited to confirmed cross-registrations considered as “Completed” in both registrations. The analysis of registration status was limited to confirmed cross-registrations with an available start date in the registry, and is only reported for ClinicalTrials.gov and DRKS (all EUCTR trials should be prospectively registered, as registration is linked to regulatory approval by national competent authorities).

We performed post-hoc sensitivity analyses to explore the robustness of our findings. In the sensitivity analysis of the completion date, completion dates in the EUCTR were taken solely from the protocol for Germany (i.e., EUCTR results pages were disregarded). For results reporting, we performed two sensitivity analyses where the definition of summary results was broadened to include 1) other reporting formats uploaded in the summary results field on EUCTR and DRKS, including synopses, summary or study reports, statements of termination, structured results reports in the format of another registry, and linked publications, 2) linked publications on ClinicalTrials.gov. Our analysis was descriptive; we did not test specific hypotheses. More detailed information is available in **Supplement S2**.

### Data and Code Availability

Data processing was performed in R 4.4.2 (20) and Python 3.9 (Python Software Foundation, Wilmington, Delaware USA). The code to identify and analyze cross-registrations is openly available on GitHub: https://github.com/maia-sh/intovalue-data and https://github.com/quest-bih/intovalue-crossreg. The data are openly available on Zenodo (https://doi.org/10.5281/zenodo.14949157).

## Results

### Identification of potential EUCTR cross-registrations

Based on registration and publication data, we identified 1,025 potential EUCTR cross-registrations in the IntoValue dataset (i.e., where each potential cross-registration involves an EUCTR registration and a ClinicalTrials.gov or DRKS registration). After applying our exclusion criteria (see Methods), this led to 625 potential EUCTR cross-registrations, of which 90% (n = 563/625) were linked to IntoValue trials registered on ClinicalTrials.gov and 10% (n = 62/625) to IntoValue trials registered on DRKS (**Figure 1**). These 625 potential cross-registrations involved 614 unique registrations from IntoValue (11 TRNs in IntoValue were each linked to 2 EUCTR TRNs) and 620 unique EUCTR registrations (5 EUCTR TRNs were each linked to more than one registration in DRKS or ClinicalTrials.gov). Thus, based on this approach, the proportion of IntoValue trials with a potential EUCTR cross-registration was 21% (n = 614), and ranged between 18% and 25% per completion year (**Supplement S3**).

**Figure 1.**
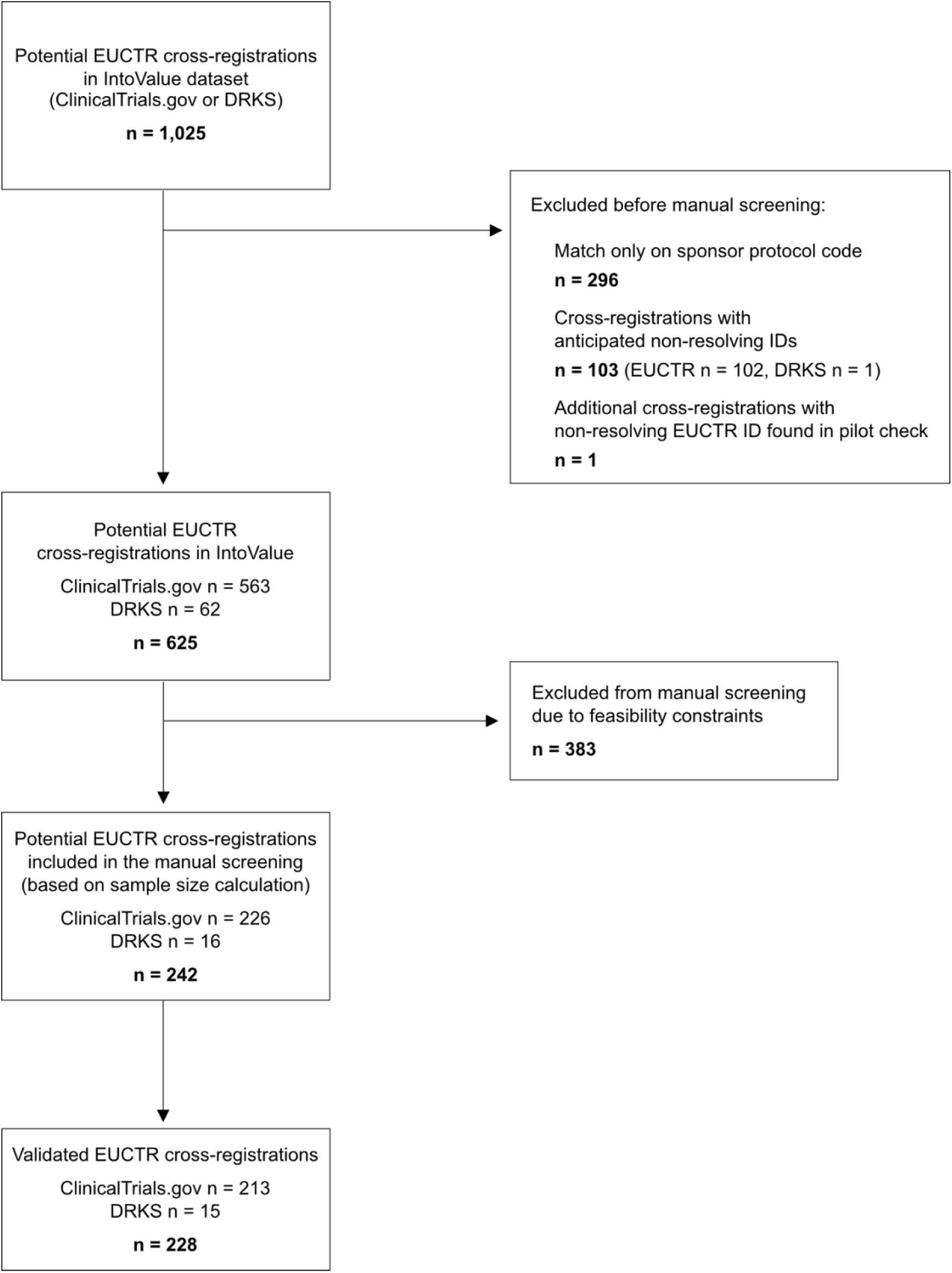
Identification and screening of potential EUCTR cross-registrations in IntoValue. Potential cross-registrations were grouped into hierarchical categories based on how they were linked (e.g., title matching). The number of cases screened in each category was determined based on sample size calculation and a prespecified stopping rule.

Potential cross-registrations were linked through a combination of factors (**Figure 2, Supplement S3**). The majority of potential cross-registrations (75%, n = 470) were linked through a TRN listed in one or both registries (alone or in combination with other factors). Only 17% (n = 109) were linked through a TRN in both registries. Matching trials based on characteristics beyond TRNs in the registry led to an additional 155 (25%) potential EUCTR cross-registrations. Of these additional cross-registrations, 70% (n = 108) matched exclusively on title (approximate match), 19% (n = 30) matched exclusively on a TRN in a related results publication, and 11% (n = 17) were linked via both factors.

**Figure 2.**
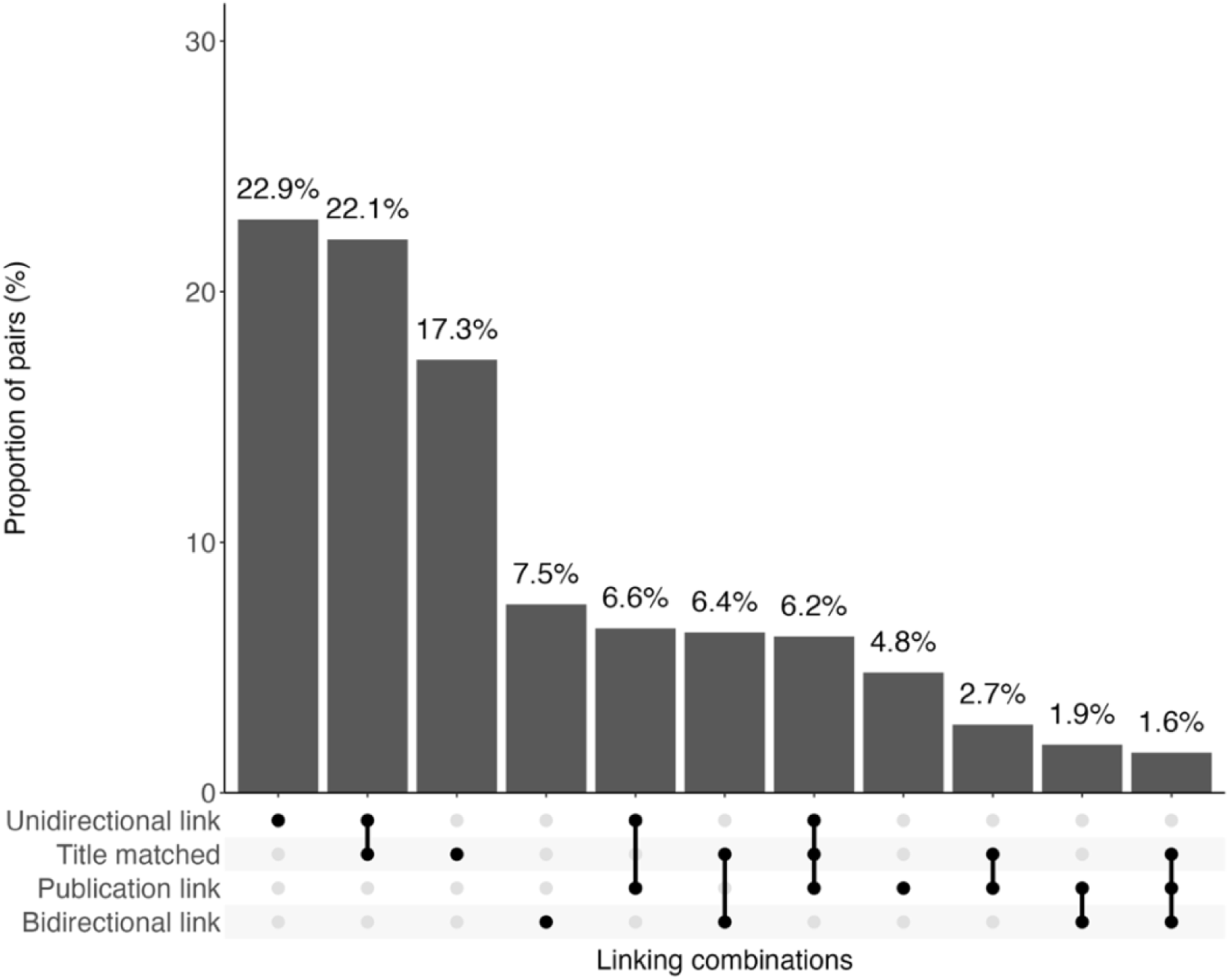
Percentage of potential EUCTR cross-registrations (n = 625) with each combination of characteristics indicating a cross-registration. Note that while bidirectional registry links inherently imply a unidirectional registry link, these were counted separately.

Focusing on potential cross-registrations linked through a TRN listed in one or both registries, EUCTR provided a matching ClinicalTrials.gov TRN for 53% (n = 216/410), and ClinicalTrials.gov provided a matching EUCTR TRN for 72% (n = 294/410). In turn, EUCTR provided a matching DRKS TRN for 15% (n = 9/60), and DRKS provided a matching EUCTR TRN for 100% (n = 60/60) (**Figure 3**).

**Figure 3.**
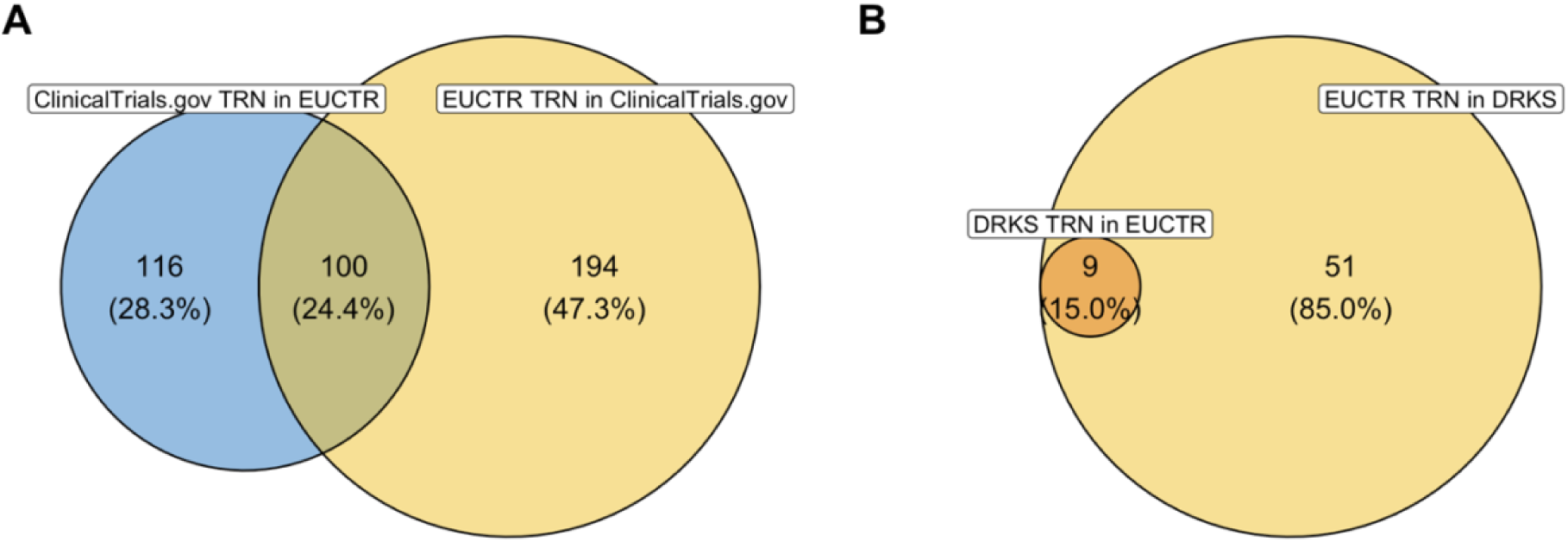
Source of TRNs for potential EUCTR cross-registrations linked through a TRN in the registry (n = 470). **A.** EUCTR – ClinicalTrials.gov, n = 410. **B.** EUCTR – DRKS, n = 60.

We queried the MDR (ECRIN) via the API to obtain secondary identifiers associated with the 2,895 trials in IntoValue. We obtained 527 unique EUCTR TRNs associated with 530 unique TRNs in IntoValue, from which we could infer 531 potential EUCTR cross-registrations. **Supplement S4** shows the overlap of potential cross-registrations found using our search strategy (n = 729) (see Methods) and based on the data obtained from the MDR query. Of the 729 potential cross-registrations identified using the approach in this study, 511 (70%) could also be identified based on the MDR query. In turn, 218 (30%) potential cross-registrations were only found through our approach. More than half of these were exclusively linked via approximate title matching or a TRN in a related publication. In turn, 20 potential cross-registrations were exclusively inferred based on the MDR query. More detailed information is available in **Supplement S4**.

### Manual validation of cross-registrations

Of the 625 potential cross-registrations identified, 39% (n = 242; EUCTR – ClinicalTrials.gov n = 226; EUCTR – DRKS n = 16) were manually screened across all categories. Of these, 94% (n = 228; EUCTR – ClinicalTrials.gov n = 213, EUCTR – DRKS n = 15) were confirmed as cross-registrations of the same trial according to the definition and criteria used in this study (**Table 1**). Of these, 6 did not have a protocol for Germany in the EUCTR. Almost all EUCTR cross-registrations identified via a TRN in one or both registries (alone or in combination with other factors) could be confirmed. The manual review led to 14 false positive cases: 9 were linked exclusively through a TRN in the full text of a related results publication, 2 were linked exclusively via a TRN in one of the registrations, and 3 were linked exclusively via approximate title matching. All 30 cross-registrations linked exclusively via a TRN in a related results publication were manually reviewed, and 21 (70%) of these could be confirmed. Of these, 16 were linked exclusively via a TRN in the publication full text. More information on screened cross-registrations and the outcome of the manual review is available in **Supplement S5**.

**Table 1.** Potential EUCTR cross-registrations that were identified, manually screened, and confirmed across categories. Categories are exclusive and hierarchical (bidirectional linking > unidirectional linking > approximate title matching > link via publication).

|  | Registry | Overall<br>n = 625 | Manually screened<br>n = 242 | Confirmed<br>n = 228 |
| --- | --- | --- | --- | --- |
| **Cross-registrations with bidirectional registry links** | **(Subtotal)** | **109** | **64** | **64 (100%)** |
|  | ClinicalTrials.gov | 100 | 59 | 59 |
|  | DRKS | 9 | 5 | 5 |
| **Remaining cross-registrations with unidirectional registry link** | **(Subtotal)** | **361** | **81** | **79 (98%)** |
|  | ClinicalTrials.gov | 310 | 71 | 69 |
|  | DRKS | 51 | 10 | 10 |
| **Remaining cross-registrations with approximate title matching** | **(Subtotal)** | **125** | **67** | **64 (96%)** |
|  | ClinicalTrials.gov | 124 | 67 | 64 |
|  | DRKS | 1 | 0 | 0 |
| **Remaining cross-registrations with match only on TRN in publication** | **(Subtotal)** | **30** | **30** | **21 (70%)** |
|  | ClinicalTrials.gov | 29 | 29 | 21 |
|  | DRKS | 1 | 1 | 0 |

### Discrepancy analysis

For confirmed cross-registrations (n = 228), we compared the following characteristics across registrations of the same trial: registration status, recruitment status, completion date, and availability of summary results in the registry. Of the 213 confirmed EUCTR – ClinicalTrials.gov registrations, 62% (n = 133) were registered prospectively in ClinicalTrials.gov. In turn, of the 15 confirmed EUCTR – DRKS cross-registrations, 73% (n = 11) were registered prospectively in DRKS (**Supplement S6**). Of confirmed registration pairs with an available recruitment status (n = 222), 85% (n = 189) had the same overall recruitment status (“Completed”) in both registry entries according to the mapping used in this study (**Supplement S2**). The remaining 15% (n = 33) had discrepant recruitment statuses across registry entries (8%, n = 17, “Completed” on EUCTR and “Other” on the other registry; 5%, n = 12, “Ongoing” on EUCTR and “Completed” on the other registry; 2%, n = 4, “Ongoing” on EUCTR and “Other” on the other registry) (**Supplement S6**).

Of the 228 manually confirmed cross-registrations, 184 had an overall recruitment status “Completed” and an available completion date in both registry entries (n = 170 EUCTR – ClinicalTrials.gov, n = 14 EUCTR – DRKS). Of these, 50% (n = 92; ClinicalTrials.gov n = 84; DRKS n = 8) had the same completion date (i.e., within the same year-month) across registrations (**Figure 4**). Focusing on discrepant cases, differences ranged from 1 – 110 months (median = 5, standard deviation = 15). The completion date in EUCTR was earlier in 36% (n = 67) and later in 14% (n = 25) of cases. The same analysis using completion dates in the EUCTR protocol for Germany (i.e., completion dates in results sections were disregarded) (n = 170) led to similar overall results, except that differences in completion dates for discrepant cases were smaller, ranging from 1 – 42 months (median = 5, standard deviation = 7) (**Supplement S6**).

**Figure 4.**
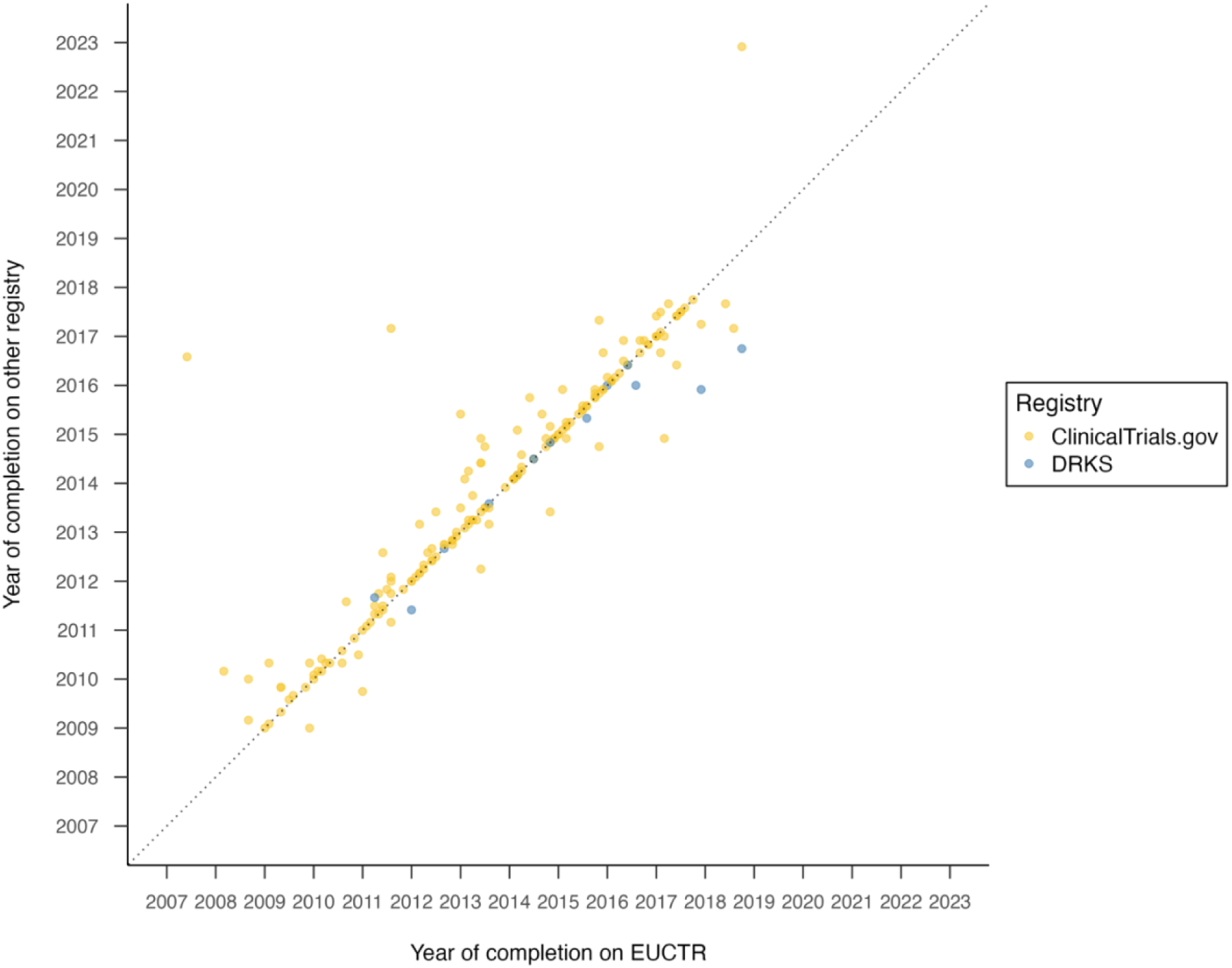
Analysis of completion dates (month/year) across registrations of the same trial. Completion dates in EUCTR were taken from the results sections (if available), otherwise from the protocol for Germa

Of the EUCTR – ClinicalTrials.gov cross-registrations considered as “Completed” in both registries (n = 181), 30% (n = 55) had only reported structured results in EUCTR, 11% (n = 20) only in ClinicalTrials.gov, 7% (n = 13) in both registries, and 51% (n = 93) had not reported structured tabular results in either registry (**Figure 5**). Broadening the definition of summary results to include other formats uploaded in the same field on EUCTR (not applicable for ClinicalTrials.gov) led to an almost two-fold increase in reporting exclusively on the EUCTR (59%, n = 107), and an overall increase in reporting from 49% to 77%. Using this definition, 15% (n = 27) of cross-registrations had posted summary results on both EUCTR and ClinicalTrials.gov (**Figure 5**). **Supplement S6** displays reporting rates additionally accounting for publications listed in the separate “Publications” section on ClinicalTrials.gov.

**Figure 5.**
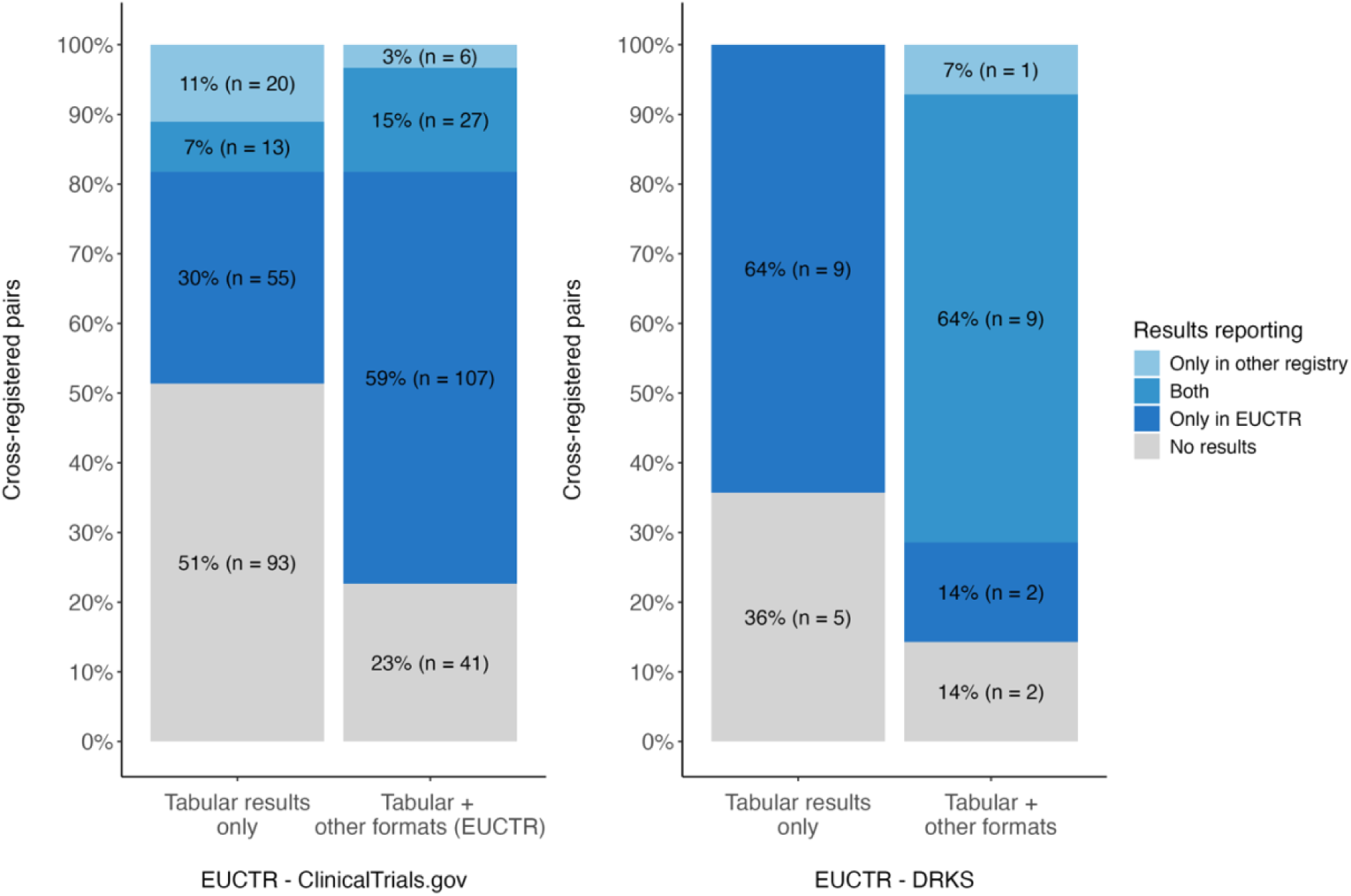
(Summary) results reporting for confirmed EUCTR – ClinicalTrials.gov (left) and EUCTR – DRKS (right) cross-registrations. Main analysis only considers structured tabular results formats; sensitivity analysis also considers other formats uploaded in the summary results field on EUCTR and DRKS.

For EUCTR – DRKS cross-registrations considered as “Completed” on both registries (n = 14), summary results reporting was 64% (n = 9) and was driven entirely by the EUCTR, since DRKS does not provide a format for structured tabular results. Broadening the definition of summary results to include other results formats uploaded in the same fields on EUCTR and DRKS (including linked publications) increased overall reporting to 86% (n = 12), with almost two thirds of cross-registrations (64%, n = 9) having reported in some form on both registry entries (**Figure 5**). More detailed information on the inclusion of reporting formats and registry fields across analyses is available in **Supplement S2**.

## Discussion

### Summary of findings

We identified and characterized EUCTR cross-registrations in an existing dataset of 2,895 clinical trials conducted at German UMCs and registered on ClinicalTrials.gov or DRKS. Using a combined approach based on registry data and trial results publications, we identified 625 potential EUCTR cross-registrations that were linked through a combination of factors. While TRNs in the registry accounted for the majority of potential cross-registrations, only 17% of cross-registrations were linked via a TRN in both registries, limiting their findability. Approximate title matching, and to a lesser extent TRN mentions in trial results publications, were effective strategies to identify additional cross-registrations. For a sample of manually confirmed EUCTR cross-registrations, 85% had the same overall recruitment status while only 50% had the same completion date across registrations of the same trial. EUCTR was the dominant route for summary results posting, and included structured tabular results as well as other formats. Overall, the availability of summary results across registrations of the same trial varied substantially depending on which reporting formats and registry fields were considered. The findings of this descriptive study indicate room for improvement in how cross-registrations are linked, and raise concerns about the reliability of information in trial registries, despite their role in promoting transparency in medical research. Our findings also illustrate the importance of considering cross-registrations for trial searches, evidence synthesis, and meta-research studies.

### Findings in context

Our findings align with previous work that cross-registration of trials is prevalent, and failure to account for cross-registrations may impact conclusions when identifying unique trials (21) or monitoring adherence to responsible research practices such as results dissemination (22). Previous studies have investigated duplicate trial records, most notably in the ICTRP as well as in individual registries. Methods to identify “hidden duplicates” using more advanced automated strategies have been proposed and include string matching (23), text-based similarity scoring (21), and random forest classifiers (17). In line with some of this work (21,23), our findings suggest that title matching is an effective strategy to identify hidden duplicates. We additionally leveraged trial results publications to identify additional cross-registrations based on TRNs in the publication abstract, secondary identifier, and full text. However, while this approach identified approximately 5% of potential cross-registrations not already linked through other factors, almost a third of these were false positives based on a TRN in the publication full text. Given the resources needed to follow up results publications associated with trials and poor linking between trial registrations and publications (24,25), this approach may only be worthwhile if trial results publications are readily available.

The lack of consistency across registrations of the same trial is broadly consistent with previous work. Fleminger and colleagues (26) found that 16.2% of 10,492 trials registered on ClinicalTrials.gov and EUCTR were discrepant on completion status, with the majority “completed” on ClinicalTrials.gov but “ongoing” on EUCTR (26). Speich et al. (2021) assessed the consistency of trial information of 197 studies registered in more than one registry, and found 46% agreement for trial status and 62% agreement for the availability of summary results in the registry (27). Another study by DeVito et al. investigating results dissemination routes of EUCTR trials found that the availability of results on EUCTR was higher than on other registries with matched records, and 14% of trials with results were only available on EUCTR (25). In the present study, EUCTR was also the dominant route for reporting summary results. This is perhaps not surprising given the legal requirement to report results in the EUCTR as well as the possibility to upload summary results in different formats (e.g., non-structured synopses).

### Strengths, Limitations, and Challenges

A strength of this project is the comprehensiveness of the approach used to identify cross-registrations, which included searching for TRNs in various registry fields and trial results publications, title matching, as well as cleaning of TRNs. The manual review of cross-registrations ensured the validity of the evaluation of discrepancies across registrations of the same trial and allowed for a nuanced description of summary results and challenges extracting information from registries. The code and data generated in this study are openly available and can be adapted for use in other studies.

This project has several limitations. Our approach builds on an initial set of potential cross-registrations downloaded on 1 November 2022 for a previous study and therefore the ClinicalTrials.gov and DRKS data used to identify cross-registrations may no longer be up to date. However, since trials in IntoValue were completed over 5 years ago, we do not expect substantial changes in the registry data. Though regular expressions and text matching allowed for non-exact matches, true cross-registrations may have been missed if TRNs were severely misformatted or if their titles differed significantly. In turn, despite our systematic approach to validate cross-registrations, we cannot exclude that some cases may have been erroneously classified as such. However, the aim of this study was to explore and describe cross-registrations, rather than assess the precision of approaches to identify cross-registrations or systematically evaluate the transparency of cross-registered trials. Due to feasibility constraints, it was not possible to manually review all cross-registrations in the dataset, which limited the number of cross-registrations included in the discrepancy review. When assessing results reporting, we checked the type of report that was posted, but we did not perform a detailed review to confirm whether these reported on the results of the trial. Finally, trials may also have been registered in duplicate in the same registry or in further registries beyond the scope of this study.

The following challenges should be noted. As reported elsewhere (17), we encountered differences in available fields and distinct terminologies across registries, making it difficult to compare characteristics across registrations of a trial. This was particularly challenging for summary results reporting, given the wide variety of formats for reporting results and differences in reporting fields across registries. The ICTRP has drafted recommendations for the minimum elements of study results to be reported in registries to promote greater harmonization (28). We encountered several edge cases while manually validating cross-registrations, many of which concerned multinational trials with different enrollment across registrations. We also came across quite a few trials that listed an EUCTR identifier, which did not resolve in the EUCTR. While this is expected in some cases (e.g., phase 1 trials conducted on adults are not publicly available on EUCTR), other cases remained unclear, which poses an additional challenge for monitoring assessments.

### Implications for policy and practice

The WHO International Standards for Clinical Trial Registries outline responsibilities for registrants, including that a trial must be registered in the fewest number of registers necessary to meet applicable laws and regulations. Furthermore, if a trial is registered on more than one registry, all identifiers for the trial should be submitted to each registry as secondary identifiers, and a single point of contact for the trial should be responsible for ensuring that the same data is provided to each registry (29). While we did not investigate whether trials in our dataset were required to be registered on more than one registry, the missing links and discrepancies in key characteristics indicate that trials fall short of these international standards.

Unambiguous identification of trials is essential to ensure the identification and inclusion of relevant evidence in systematic reviews and meta-analyses, which form the basis of clinical guidelines.

Unambiguous identification of trials is also important in the context of audits of trials’ adherence to transparency standards, e.g., reporting of trial results. For instance, a cross-registered trial that reported results in one registry but not in the other may appear unreported if the records are not linked or if only a single registry is examined. Even if clearly linked, cross-registered records raise questions for monitoring adherence to transparency standards, such as what qualifies as adherence. A trial registered on both DRKS and EUCTR with results only in EUCTR may well be considered as “reported” in an audit. Yet, a patient may still face challenges finding the results of the trial.

Inconsistent information across registrations of the same trial also complicates studies of the clinical research enterprise. Studies assessing the timeliness of results reporting typically use the (primary) completion date on the registry to determine whether trial results are due (4). In the case of discrepant completion dates, which we observed in 50% of confirmed cross-registrations, a trial may seem due to report in one registry but not in the other. Differences in available fields and formats across registries also complicate how to monitor practices. In EUCTR, results were posted as structured results as well as a range of other formats; DRKS did not provide a structured tabular results format directly on the registry, and results reporting fields could include linked publications or reports; in turn, ClinicalTrials.gov provides separate sections for structured summary results and publications, either manually linked in the registry or automatically linked via PubMed. This raises the challenge of how to effectively communicate trial adherence without losing important nuance relating to the contents of the registration. Finding appropriate ways to identify, handle, and describe cross-registrations is particularly relevant for studies conducted in jurisdictions such as Germany, where trials are often registered on more than one registry. Previous audits of trial transparency involving multiple registries have either de-duplicated trials in favor of a specific registry (3) or merged registrations to use the most updated information (5,22).

### Actionable areas for stakeholders

We reiterate previous calls to minimize cross-registrations unless absolutely required, and should a trial be registered on more than one registry, to provide TRNs in all registry records and trial results publications, and ensure that all registrations of a trial are accurate and up to date (23,26,29).

Complementary to existing recommendations, our findings suggest that the WHO joint statement on public disclosure of results from clinical trials (6) may benefit from further guidance on results reporting for cross-registered trials. While all records of the same trial should ideally be maintained and consistent to increase trials’ usefulness, in practice this requires additional coordination from trialists. Thus, efforts to further harmonize registries and requirements, as well as reduce burden on researchers may be fruitful.

When searching for cross-registrations, we encountered misformatted TRNs that limit findability. Registries could facilitate the identification of cross-registrations by validating the format of TRNs provided in registrations, and flagging trials that are likely cross-registered (e.g., drug trials registered on DRKS or ClinicalTrials.gov with enrollment in the EU, which by law must be registered on EUCTR or CTIS). Trial records that have been imported from one registry to another should be clearly labelled to differentiate these from intentional registrations from trialists, and updates to the source registration should be reflected in the imported registration. Further development of infrastructures that aggregate and process information across data sources (e.g., ICTRP, ECRIN MDR) could present an opportunity to highlight inconsistences across records and prompt trialists to update their registrations. Using the MDR API, we successfully identified approximately 70% of potential cross-registrations detected by our approach, highlighting its potential to efficiently flag potential cross-registrations at scale. Importantly, these platforms should be designed such that queries can be executed programmatically, enabling regular updates and integration with automated monitoring workflows and tools (30).

### Future research directions

Future studies may consider examining how cross-registrations are linked across other registries. Our exploratory analysis of the reliability of trial information in a small number of trials registered on both EUCTR and DRKS suggests that a more detailed characterization of results reporting in DRKS may be warranted, particularly given recent changes to the registry. Moreover, a better understanding of the circumstances that lead to a trial being registered on more than one registry would inform whether to focus improvement efforts on minimizing unnecessary cross-registration, or ensuring the consistency of registrations of the same trial, or both. Future work may also consider identifying gaps or discrepancies in guidance provided by research institutions and funders in relation to WHO standards (29), and whether and how these stakeholders consider cross-registrations in their policies and internal monitoring assessments.

## Conclusions

Despite WHO standards that promote the unambiguous identification of clinical trials, registrations of the same trial are often inadequately linked and present inconsistent information. Our findings suggest the need for additional guidance and coordination on cross-registration and highlight the importance of considering cross-registrations in evidence synthesis and studies of the clinical research enterprise.

## Author Contributions

**LF:** Conceptualization, Data curation, Formal analysis, Investigation, Methodology, Software, Visualization, and Writing - review & editing. **EBA:** Conceptualization, Data curation, Formal analysis, Investigation, Methodology, Software, Validation, Visualization, and Writing - review & editing. **SY:** Conceptualization, Methodology, Validation, and Writing - review & editing. **MSH:** Conceptualization, Data curation, Methodology, Resources, Software, Validation, Visualization, and Writing - review & editing. **VN:** Software, Validation, Visualization, and Writing - review & editing. **SGS:** Conceptualization, Methodology, Validation, and Writing - review & editing. **DS:** Conceptualization, Methodology, Project administration, and Writing - review & editing. **DLF:** Conceptualization, Data curation, Investigation, Methodology, Project administration, Software, Supervision, Validation, Visualization, Writing - original draft, and Writing - review & editing.

## Data Availability

Data processing was performed in R 4.4.2 and Python 3.9 (Python Software Foundation, Wilmington, Delaware USA). The code to identify and analyze cross-registrations is openly available on GitHub: https://github.com/maia-sh/intovalue-data and https://github.com/quest-bih/intovalue-crossreg. The data are openly available on Zenodo (https://doi.org/10.5281/zenodo.14949157).

https://github.com/maia-sh/intovalue-data

https://github.com/quest-bih/intovalue-crossreg

## Acknowledgements

We thank Nick DeVito for providing some of the registry data used in this study. We thank Merle Marie-Pittelkow and Anna Lene Seidler for their input on the protocol.

