## Supplemental Materials for "Accounting for cross-registration in monitoring responsible research in clinical trials: A cross-sectional study of trials at German university medical centers"

### Supplements

#### Supplement S1: Approach to identify potential cross-registrations

We used the following data sources to identify potential EUCTR cross-registrations in IntoValue:

1. IntoValue dataset, which lists all Trial Registration Numbers (TRNs) in the IntoValue 1 and IntoValue 2 cohorts (`trials.csv`, https://github.com/maia-sh/intovalue-data/tree/main/data/processed). Note that an additional filtering step is required to limit the `trials.csv` dataset to trials that meet the IntoValue inclusion criteria (i.e., interventional, completion date between 2009 – 2017, and associated with a German University Medical Center (UMC) as lead sponsor, responsible party, or with a principal investigator from a UMC). Furthermore, duplicate trials were de-duplicated in favour of the trial data from IntoValue 2. Applying these inclusion criteria led to a dataset of 2,895 trials, which formed the basis for this study. This dataset is referred to as **"IntoValue trials"** in **Figure S1-1**.
2. Identifiers listed on ClinicalTrials.gov and DRKS registrations associated with trials in IntoValue (download date: 1 November 2022). Identifiers include TRNs, grant numbers, and sponsor codes. Identifiers obtained from ClinicalTrials.gov and DRKS were listed in separate tables, referred to in **Figure S1-1** as **"IntoValue: Additional identifiers listed in ClinicalTrials.gov"** and **"IntoValue: Additional identifiers listed in DRKS"**, respectively.
3. Existing dataset of potential cross-registrations in IntoValue based on a) TRNs listed in ClinicalTrials.gov and DRKS and b) TRNs mentioned in the abstract, secondary identifier, or full text of associated results publications (`cross-registrations.rds`, https://github.com/maia-sh/intovalue-data/tree/main/data/processed/trn). This dataset is referred to in **Figure S1-1** as **"IntoValue: Potential cross-registrations based on registry and publications"**.
4. Full EUCTR data, which includes all protocols (download date: 3 February 2024) and all results (download date: 15 February 2024). EUCTR protocols and results are referred to in **Figure S1-1** as **"EUCTR protocols"** and **"EUCTR results"**, respectively. TRNs were extracted and cleaned from both the EUCTR protocols (all country protocols) and results (if available). Cleaning of TRNs was done partially with the {ctregistries} R package (https://github.com/maia-sh/ctregistries) and through custom-made regular expressions.

**Figure S1-1** displays the data sources and data processing steps to identify potential cross-registrations in the IntoValue dataset. One of the processing steps involved approximate title matching between IntoValue trials registered on ClinicalTrials.gov (official title) or DRKS and trials registered on the EUCTR with "Germany" listed as a member state (full title). We quantified the dissimilarity between titles using the optimal string alignment (osa) method of the amatch function in the {stringdist} R library (1). The dissimilarity metric was therefore the restricted Damerau-Levenshtein distance, which is the number of deletions, insertions, substitutions, and adjacent character transpositions to convert one string of characters to another (1). The argument maxDist of the amatch function was set to 10 (informed by pilot manual checks).

In the final step, the three resulting tables were combined into a single dataset capturing a unique cross-registration pair per row and the ways in which each pair was connected. Each potential cross-registration pair was then assigned a priority according to its level of connection. A filtered version of the final dataset was used as basis for the manual validation of cross-registrations and all other analyses. More detailed information is openly available in GitHub: https://github.com/maia-sh/intovalue-data and https://github.com/quest-bih/intovalue-crossreg. Screenshots displaying where identifiers were extracted from the registries are included at the end of this Supplement.

**Figure S1-1.** Overview of the approach to identify potential EUCTR cross-registrations in the IntoValue dataset. The approach combined existing and new data sources, and was based on title matching (green), TRNs in trial results publications (yellow), and identifiers in the registry (light blue).

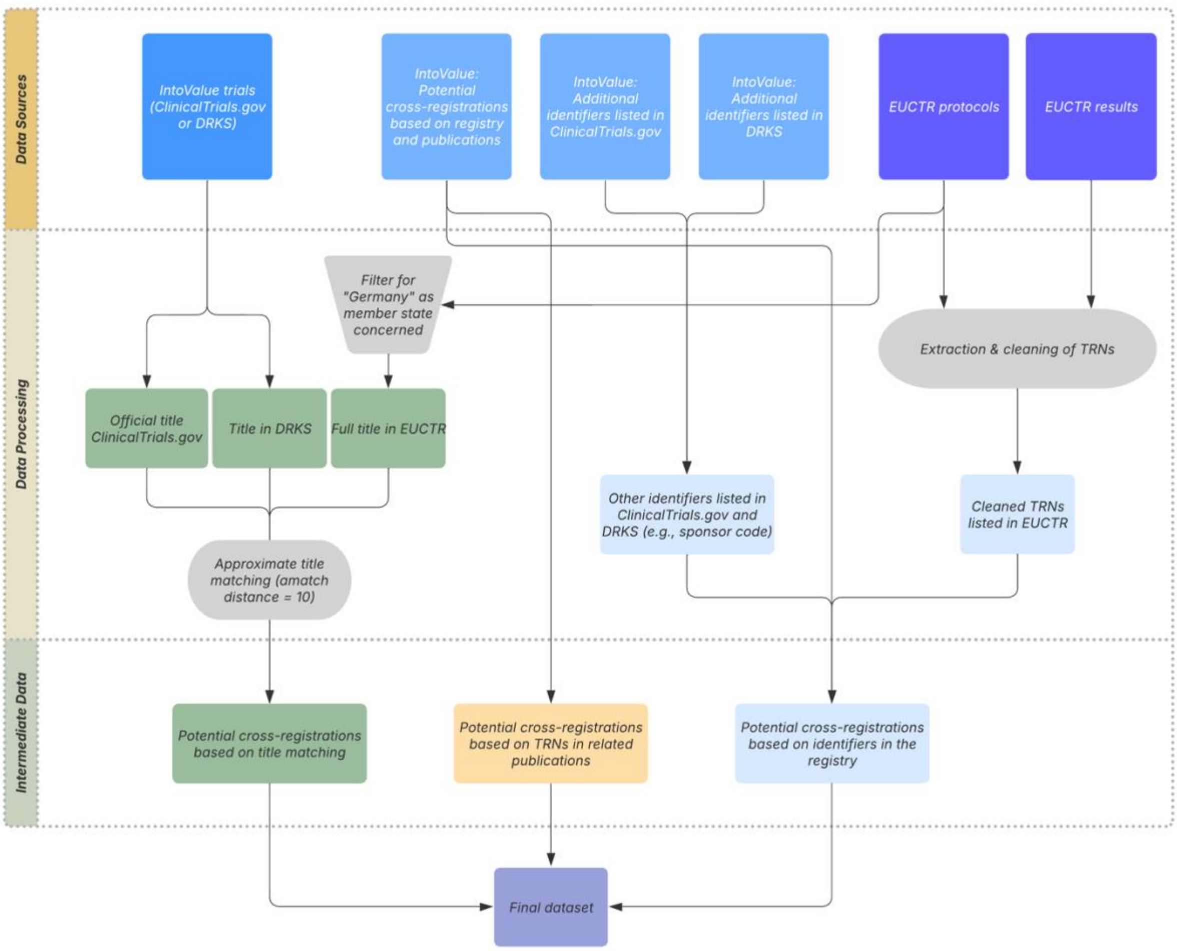

#### Examples of where identifiers were extracted from registries

##### ClinicalTrials.gov

**Figure S1-2.** Example EUCTR TRN listed in a ClinicalTrials.gov registration. In this example, the EUCTR TRN is mis-formatted.

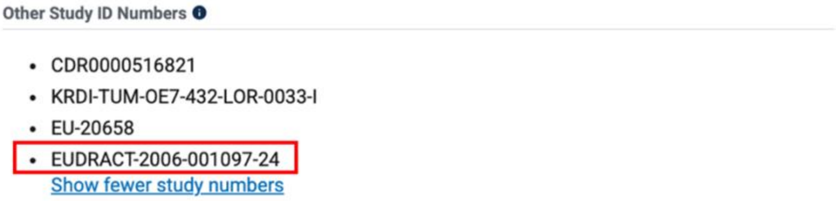
Other Study ID Numbers

- CDR0000516821
- KRDI-TUM-OE7-432-LOR-0033-I
- EU-20658
- EUDRACT-2006-001097-24

Show fewer study numbers

##### EUCTR

For EUCTR, TRNs and other identifiers were extracted from protocols (all country protocols) and the results (if available). This is illustrated in the screenshots below (different EUCTR trials are shown).

**Figure S1-3.** ClinicalTrials.gov TRN and sponsor protocol code listed in an EUCTR **protocol**.

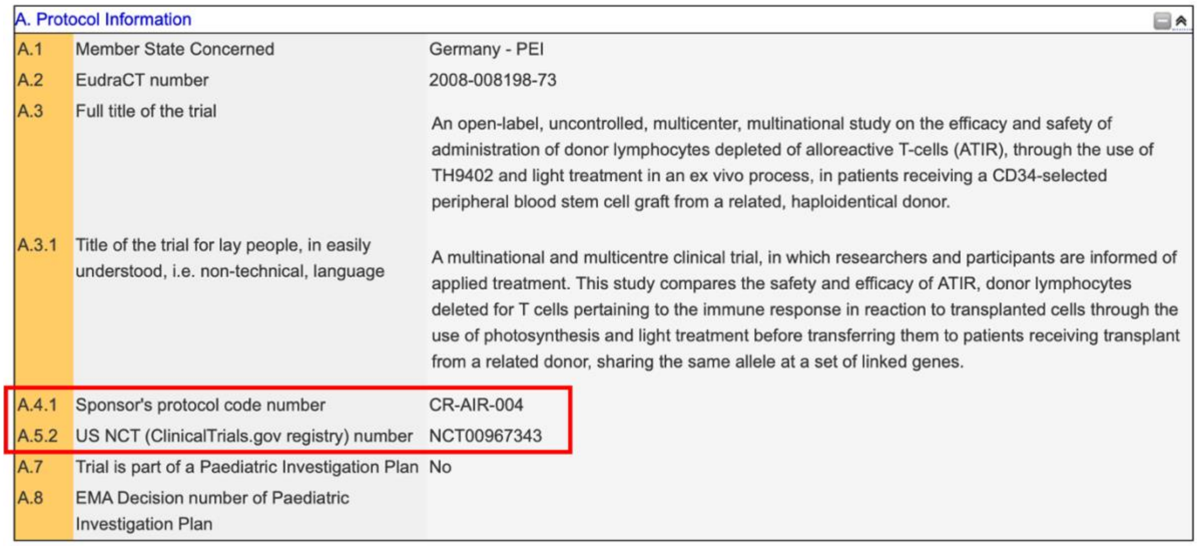
A. Protocol Information

| | | |
|---|---|---|
| A.1 | Member State Concerned | Germany - PEI |
| A.2 | EudraCT number | 2008-008198-73 |
| A.3 | Full title of the trial | An open-label, uncontrolled, multicenter, multinational study on the efficacy and safety of administration of donor lymphocytes depleted of alloreactive T-cells (ATIR), through the use of TH9402 and light treatment in an ex vivo process, in patients receiving a CD34-selected peripheral blood stem cell graft from a related, haploidentical donor. |
| A.3.1 | Title of the trial for lay people, in easily understood, i.e. non-technical, language | A multinational and multicentre clinical trial, in which researchers and participants are informed of applied treatment. This study compares the safety and efficacy of ATIR, donor lymphocytes deleted for T cells pertaining to the immune response in reaction to transplanted cells through the use of photosynthesis and light treatment before transferring them to patients receiving transplant from a related donor, sharing the same allele at a set of linked genes. |
| A.4.1 | Sponsor's protocol code number | CR-AIR-004 |
| A.5.2 | US NCT (ClinicalTrials.gov registry) number | NCT00967343 |
| A.7 | Trial is part of a Paediatric Investigation Plan | No |
| A.8 | EMA Decision number of Paediatric Investigation Plan | |

**Figure S1-4.** ClinicalTrials.gov TRN and sponsor protocol code listed in the **results** of a different EUCTR trial registration.

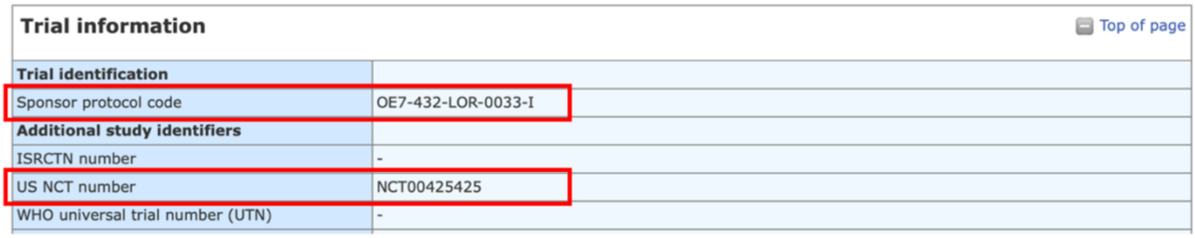
Trial information

Top of page

| | |
|---|---|
| **Trial identification** | |
| Sponsor protocol code | OE7-432-LOR-0033-I |
| **Additional study identifiers** | |
| ISRCTN number | - |
| US NCT number | NCT00425425 |
| WHO universal trial number (UTN) | - |

#### DRKS

**Fig S1-5.** EUCTR TRN listed in a DRKS registration.

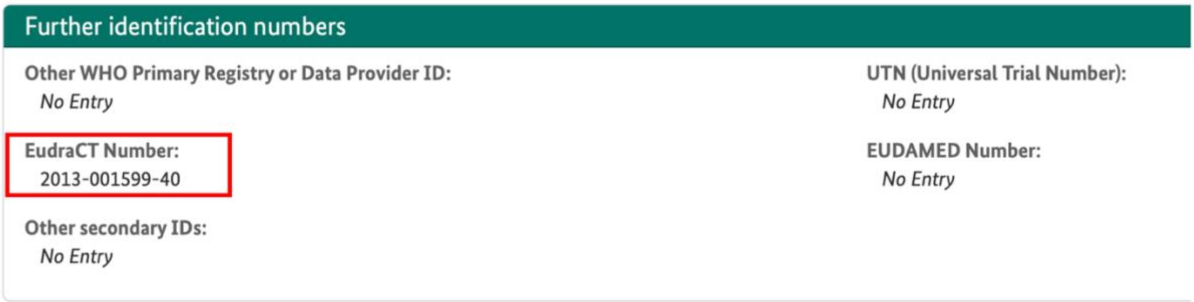


#### Non-resolving identifiers

In the pilot manual review, we encountered several potential cross-registrations involving a Trial Registration Number (TRN) that did not resolve on the registry and could therefore not be validated; these cases were removed and replaced with a new potential cross-registration to validate.

To identify additional non-resolving registrations in our dataset, we cross-checked identifiers in our dataset with lists of known registrations (full EUCTR protocol data). We also anticipated that some registrations may no longer resolve in DRKS: DRKS automatically imported studies registered on ClinicalTrials.gov with a recruitment site in Germany, however these were subsequently removed from the DRKS database with the transition to the new DRKS application at the end of 2022. See more information in the FAQ on DRKS (section *"Where do I find studies which were imported from ClinicalTrials.gov until 2017?"*). Thus, we also cross-checked identifiers in our dataset with known DRKS identifiers with non-resolving registrations. This led to 103 potential cross-registrations that involved a TRN we expected would not resolve in the registry; these were excluded from further manual validation. One additional non-resolving case found in the manual pilot and not captured with this approach was also excluded from further analysis.

Non-resolving TRNs in the EUCTR may for instance occur for phase 1 trials conducted solely on adults and that are not part of an agreed paediatric investigation plan (they are not publicly available), or due to data quality issues in the registry (2).

Previous work has also found a small proportion of non-existent ClinicalTrials.gov identifiers in clinical trial abstracts indexed by PubMed, likely due to data entry mistakes (3).

#### Supplement S2: Extraction of variables for the discrepancy review

This supplement builds on the study protocol (https://osf.io/yzxf3) and includes additional information on the assessment of discrepancies in completion date, recruitment status, and the availability of summary results. Unless specified, the registry variables were extracted from EUCTR, ClinicalTrials.gov, and DRKS per the study protocol.

##### Recruitment status

Building on Fleminger et al. (2018) (4), we mapped the recruitment status available in ClinicalTrials.gov, DRKS and EUCTR to one of the following overall recruitment statuses: “Ongoing”, “Completed”, or “Other” (see **Table S2-1**). In contrast to Fleminger et al., (2018) which considered an EUCTR trial as completed only if *all* member state protocols were either "Completed" or "Prematurely Ended", we checked the recruitment status of only the EUCTR member state protocol for Germany (if the protocol for Germany was missing, the registration pair was excluded from this analysis). The recruitment status of other member state protocols was not taken into account. Additionally, the availability of results was not an additional criterion for classifying a trial as completed, as was done by Fleminger et al. (2018) (4).

**Table S2-1.** Mapping of recruitment statuses in ClinicalTrials.gov, DRKS, and the EUCTR to allow for comparison.

| | **Ongoing** | **Completed** | **Other** |
|---|---|---|---|
| **ClinicalTrials.gov** | ‘Active, not recruiting’, ‘Available’, ‘Enrolling by invitation’, ‘Not yet recruiting’, ‘Recruiting’ or ‘Suspended’ | ‘Completed’ or ‘Terminated’ | ‘Withdrawn’, ‘Approved for marketing’, ‘No longer available’, ‘Temporarily not available’, ‘Withheld’ or ‘Unknown status’ |
| **DRKS** | ‘Recruiting planned’, ‘Recruiting ongoing’, ‘Enrolling by invitation’, ‘Recruiting suspended on temporary hold’, ‘Recruiting complete, study continuing’ | ‘Recruiting complete, study complete’ or ‘Recruiting stopped (after recruiting started)’ | ‘Recruiting withdrawn (before recruiting started)’ |
| **EUCTR** | ‘Ongoing’, ‘Restarted’, ‘Suspended by CA’ or ‘Temporarily Halted’ | ‘Completed’ or ‘Prematurely Ended’ | ‘Not Authorised’ or ‘Prohibited by CA’ |

##### Summary results

During data collection, we encountered a range of formats and registry fields used for results reporting across ClinicalTrials.gov, DRKS, and EUCTR. Therefore, we recorded the field and format for how results were reported across registries. As a deviation to the protocol, only structured tabular results in the format of the host registry and available directly on the registry were considered as "results posted" in the main analysis. This was done to highlight structured results reports, which have been shown to present more complete data on results and adverse events than journal publications (5,6). However, we note that DRKS did not provide a structured tabular results format directly on the registry at the time of the study.

We performed two post-hoc sensitivity analyses. In the first sensitivity analysis, the definition of summary results was broadened to include other formats uploaded in the summary results reporting fields on EUCTR ("summary report(s)" field) and DRKS ("Publication of study results" and "Basic reporting" fields). This could include summary reports, but also linked publications or citations (**Table S2-2**). In contrast to EUCTR and DRKS, summary results on ClinicalTrials.gov are exclusively available in the structured tabular format; in turn, linked publications are listed in a separate field. To reflect these registry differences, we performed a second sensitivity analysis to capture linked publications in ClinicalTrials.gov. When assessing results reporting, we checked the type of report that was posted, but we did not perform a detailed review of linked or uploaded documents to confirm whether they reported on the results of the trial. **Table S2-2** provides an overview of the formats we encountered (with examples) and their inclusion across analyses.

**Table S2-2.** Overview of results formats found in the EUCTR, ClinicalTrials.gov, and DRKS and their inclusion across analyses.

| Registry | Format | Example | Considered as "results posted" in the main analysis | Considered as "results posted" in first sensitivity analysis | Considered as "results posted" in second sensitivity analysis |
|---|---|---|---|---|---|
| EUCTR | Structured, tabular results | https://www.clinicaltrialsregister.eu/ctr-search/trial/2010-023688-16/results | yes | yes | yes |
| | Synopsis, study report, or summary report in the "summary report(s)" field | https://www.clinicaltrialsregister.eu/ctr-search/trial/2007-004884-24/results | no | yes | yes |
| | Tabular results from another registry in the "summary report(s)" field | https://www.clinicaltrialsregister.eu/ctr-search/trial/2007-003508-36/results | no | yes | yes |
| | Statement regarding termination of trial in the | https://www.clinicaltrialsregister.eu/ctr-search/trial/2011 | no | yes | yes |

| | | | | | |
|---|---|---|---|---|---|
| | "summary report(s)" field | -004787-30/results | | | |
| | Uploaded a file including a publication citation in the "summary report(s)" field | https://www.clinicaltrialsregister.eu/ctr-search/trial/2009-017328-25/results | no | yes | yes |
| | Uploaded abstract or paper in the "summary report(s)" field | https://www.clinicaltrialsregister.eu/ctr-search/trial/2009-012449-48/results | no | yes | yes |
| | Non-resolving link in the "summary report(s)" field | https://www.clinicaltrialsregister.eu/ctr-search/trial/2004-005223-18/results | no | no | no |
| | No results posted | https://www.clinicaltrialsregister.eu/ctr-search/search?query=2007-004333-42 | no | no | no |
| ClinicalTrials.gov | Structured, tabular results in the "Results Overview" section | https://clinicaltrials.gov/study/NCT01703819?term=NCT01703819&rank=1&tab=results | yes | yes | yes |
| | Link to publication in "Publications" section | https://clinicaltrials.gov/study/NCT00894569?term=NCT00894569&rank=1&tab=results | no | no | yes |
| | '*Results submitted, Not Posted*' in the "Results Overview" section | https://clinicaltrials.gov/study/NCT01968239?term=NCT01968239&rank=1&tab=results | no | no | no |
| | No results posted in either the "Results Overview" or "Publications" section | https://clinicaltrials.gov/study/NCT01303250?term=NCT01303250&rank=1&tab=results | no | no | no |
| DRKS | Summary results report in the "Publications/study results" field* | https://drks.de/search/en/trial/DRKS00000156 | no | yes | yes |
| | Tabular results from another registry in the "Publications/study results" field* | https://drks.de/search/en/trial/DRKS00003324 | no | yes | yes |
| | Link to publication or citation in | https://drks.de/search/en/trial/DRKS00000582 | no | yes | yes |

| | | | | | |
|---|---|---|---|---|---|
| | “Publications/study results” field* | | | | |
| | No results posted | https://www.drks.de/search/de/trial/DRKS00003217 | no | no | no |

**DRKS also includes a section entitled “Basic reporting” which includes the fields “Basic Reporting / Results tables” and “Brief summary of results”. However, in the manual review, results were almost always only found in the “Publication of study results” section, regardless of the format. We only came across one case (DRKS00009396, not a true cross-registration for 2011-001779-38) where both fields in DRKS (“Basic reporting” and “Publication of study results”) linked to the same publication.*

**Completion date**

We report no deviations from the protocol in how the data was collected for the main analysis. We additionally conducted a post-hoc sensitivity analysis where completion dates from the EUCTR were solely taken from the protocol for Germany (EUCTR registrations with no protocol for Germany or that did not have an available completion date were excluded).

### Supplement S3: Additional analyses of potential cross-registrations

The IntoValue dataset includes 2,895 trials registered on ClinicalTrials.gov (n = 2,253) or DRKS (n = 642). Using our approach, we identified 625 potential EUCTR cross-registrations. Of these, 90% (n = 563/625) were between EUCTR and ClinicalTrials.gov and 10% (n = 62/625) between EUCTR and DRKS. The 625 potential EUCTR cross-registrations involved 614 unique trials from IntoValue. Thus, the proportion of IntoValue trials with a potential EUCTR cross-registration was 21% (n = 614), and ranged between 18% and 25% per completion year (**Figure S3-1**). Of IntoValue trials registered on ClinicalTrials.gov (n = 2,253), 25% (n = 552) had a potential EUCTR cross-registration. Of the IntoValue trials registered on DRKS (n = 642), 10% (n = 62) had a potential EUCTR cross-registration.

**Figure S3-1.** Proportion of IntoValue trials with a potential EUCTR cross-registration per completion year (completion year from ClinicalTrials.gov or DRKS).

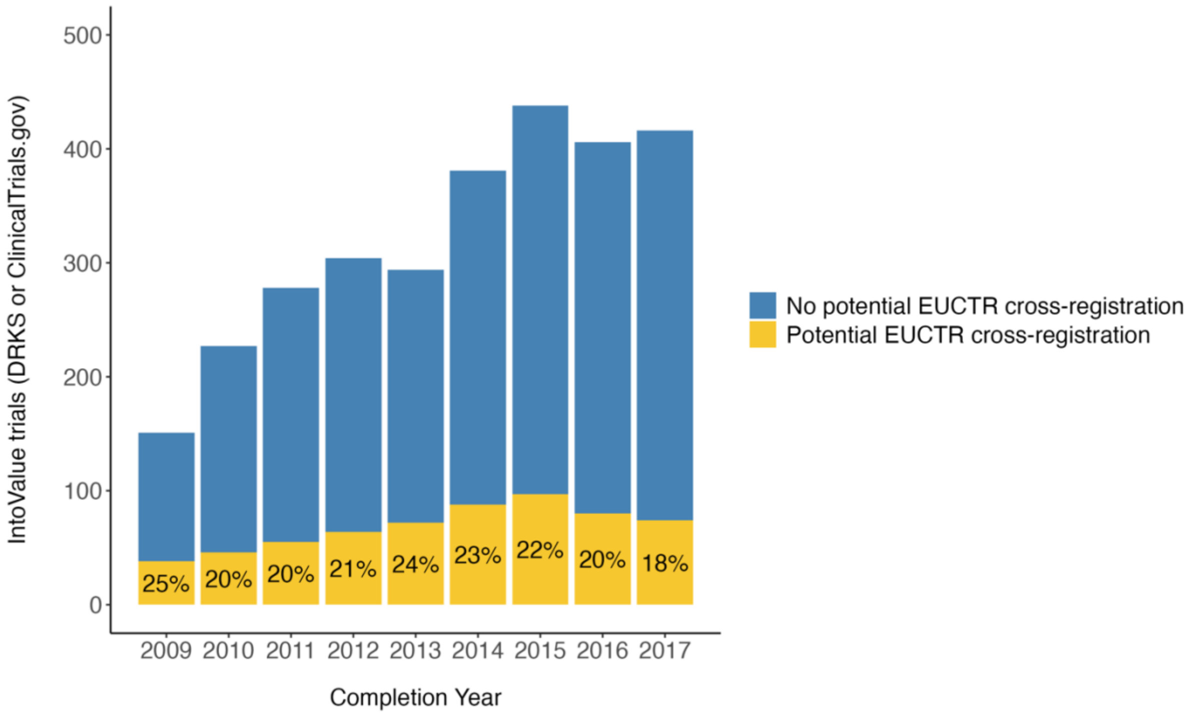

**Table S3-1** shows characteristics indicating a cross-registration (overall and by registry) for all potential EUCTR cross-registrations identifed in IntoValue (n = 625). Note that a potential cross-registration may be counted in more than one category, except for bidirectional and unidirectional linking which are exclusive, i.e., a trial pair can only ever be labelled as one of these.

**Table S3-1.** Characteristics of potential EUCTR cross-registrations in IntoValue, overall and by registry (prior to manual screening).

| | **Overall**, N = 625 | **ClinicalTrials.gov**, N = 563 | **DRKS**, N = 62 |
|---|---|---|---|
| **Bidirectional linking in registry*** | 109 (17%) | 100 (18%) | 9 (15%) |
| **Unidirectional linking in registry**** | 361 (58%) | 310 (55%) | 51 (82%) |
| **Approximate title matching** | 352 (56%) | 324 (58%) | 28 (45%) |
| **Match on identifier in publication** | 149 (24%) | 135 (24%) | 14 (23%) |

**Bidirectional linking means that both registry entries list the TRN of the registration in the other registry.*

***Unidirectional linking means that only one of the registrations lists the TRN of the other registration (the table does not specify which registry provides this information). For example, for a potential cross-registration between EUCTR and DRKS, it would imply that either the EUCTR registration mentions the TRN of the corresponding DRKS registration, or the DRKS registration mentions the TRN of the corresponding EUCTR registration.*

### Supplement S4: Comparison with ECRIN MDR

We queried the Clinical Research Metadata Repository (MDR) of the European Clinical Research Infrastructure Network (ECRIN) (https://ecrin.org/clinical-research-metadata-repository) via its API to obtain secondary identifiers associated with the 2,895 trials in IntoValue. We obtained 527 unique EUCTR TRNs associated with 530 unique TRNs in IntoValue, from which we could infer 531 potential EUCTR cross-registrations. **Figure S4-1** shows the overlap of potential cross-registrations found using our search strategy (n = 729) and based on the data obtained from the MDR query. Note that for this comparison, we excluded all potential cross-registrations identified exclusively via a matching sponsor protocol code but included potential cross-registrations involving a non-resolving TRN, since we also expected these to be listed in the MDR.

Of the 729 potential EUCTR cross-registrations we identified in IntoValue, 511 were also identified via the MDR query. In turn, 218 potential cross-registrations were only found using our approach, while 20 potential cross-registrations were only inferred based on the MDR query. An exploratory analysis suggests that the majority of the 218 potential cross-registrations found uniquely via our approach were due to the use of title matching and data sources beyond registry entries, and the fact that we also considered identifiers in the results pages as well as in the “Other identifiers” field of EUCTR entries. As for the 20 linked trial registrations exclusively found with the MDR query, while many of these registrations shared sponsor code numbers, this did not appear to account for all cases. A more detailed investigation (beyond the scope of this study) would be required to draw firmer conclusions.

**Figure S4-1.** Overlap of potential EUCTR cross-registrations obtained with our search strategy and by querying the MDR API for secondary identifiers associated with IntoValue trials.

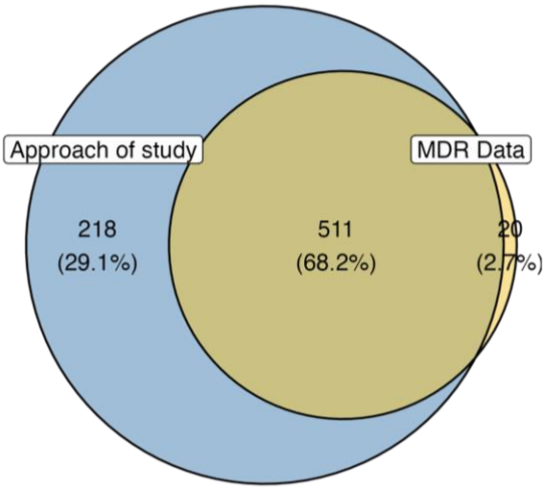

**Figure S4-2** displays an UpSet plot of potential EUCTR cross-registrations identified exclusively via the approach used in this study (n = 218) and how they were linked. Note that while bidirectional registry links inherently imply a unidirectional registry link, these were counted separately.

**Figure S4-2.** Potential EUCTR cross-registrations identified exclusively with the approach used in this study (n = 218), with each combination of characteristics indicating a cross-registration.

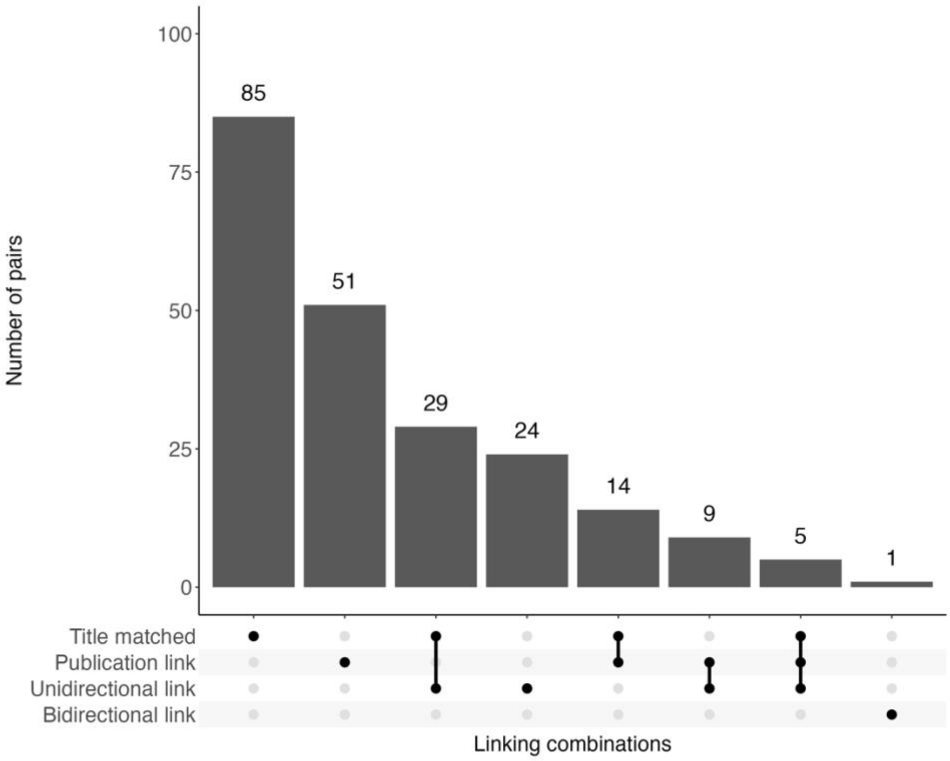

#### Supplement S5: Manual screening of cross-registrations

**Figure S5-1** displays an UpSet plot of all screened EUCTR cross-registrations (n = 242) and how they were linked, as well as the outcome of the manual screening. Note that the sample of cross-registrations included in the manual review was not drawn randomly across the entire dataset, but was informed by how cross-registrations were linked (as we expected this to influence the likelihood of finding a true cross-registration). Thus, we grouped potential cross-registrations into hierarchical categories based on how they were linked, and used sample size calculation and a prespecified stopping rule informed by the pilot manual review to determine the number of cases to manually review in each category. Cross-registrations linked exclusively via a TRN in a related results publication were all manually screened due to lower precision (hence these are overrepresented in the plot). More detailed information is available in the protocol (https://osf.io/yzxf3).

**Figure S5-1.** Manually screened cross-registrations (n = 242) with each combination of characteristics indicating a cross-registration and the outcome. Note that while bidirectional registry links inherently imply a unidirectional registry link, these were counted separately.

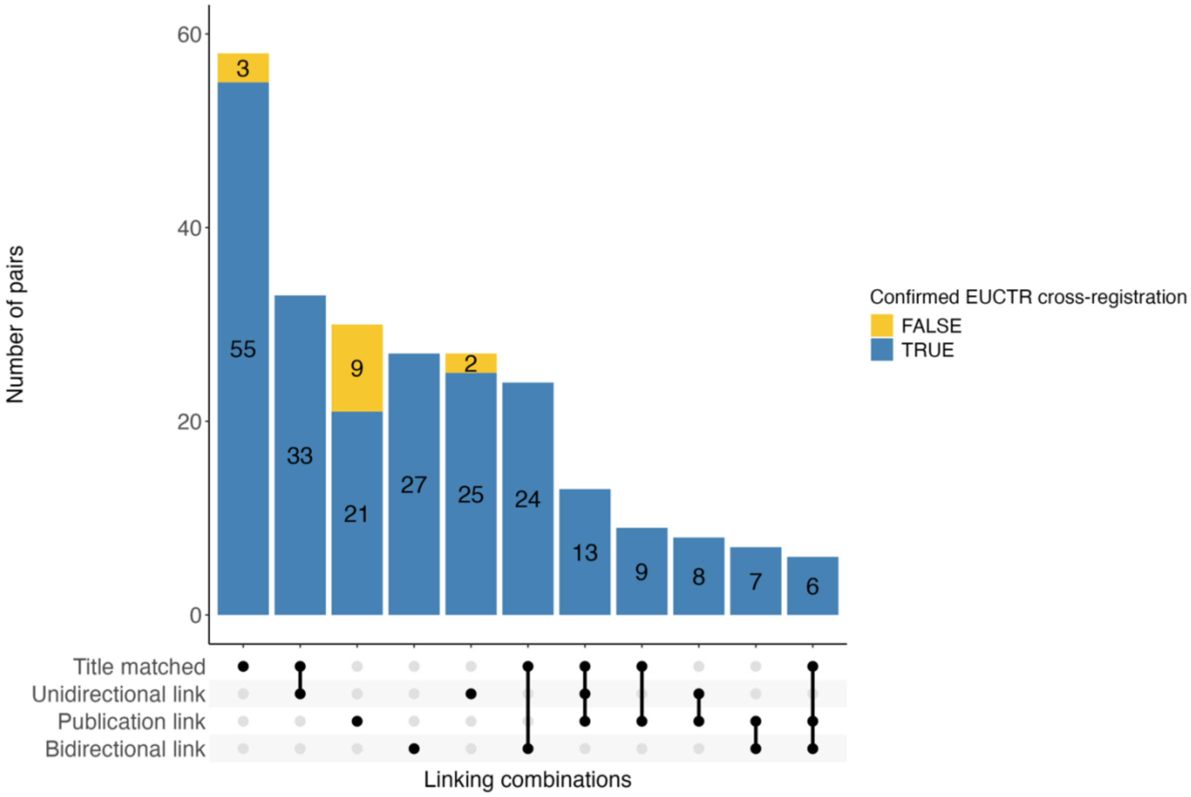

**Figure S5-2** displays an UpSet plot of screened cross-registrations that were linked via a TRN in a related results publication (alone or in combination with other factors) (n = 73). The 9 false positive cases identified through a related results publication were linked exclusively via a TRN in the full text.

**Figure S5-2.** Screened cross-registrations identified via a TRN in a related results publication (alone or in combination with other factors) with each combination of TRN location (full text, abstract, or secondary identifier) and the outcome.

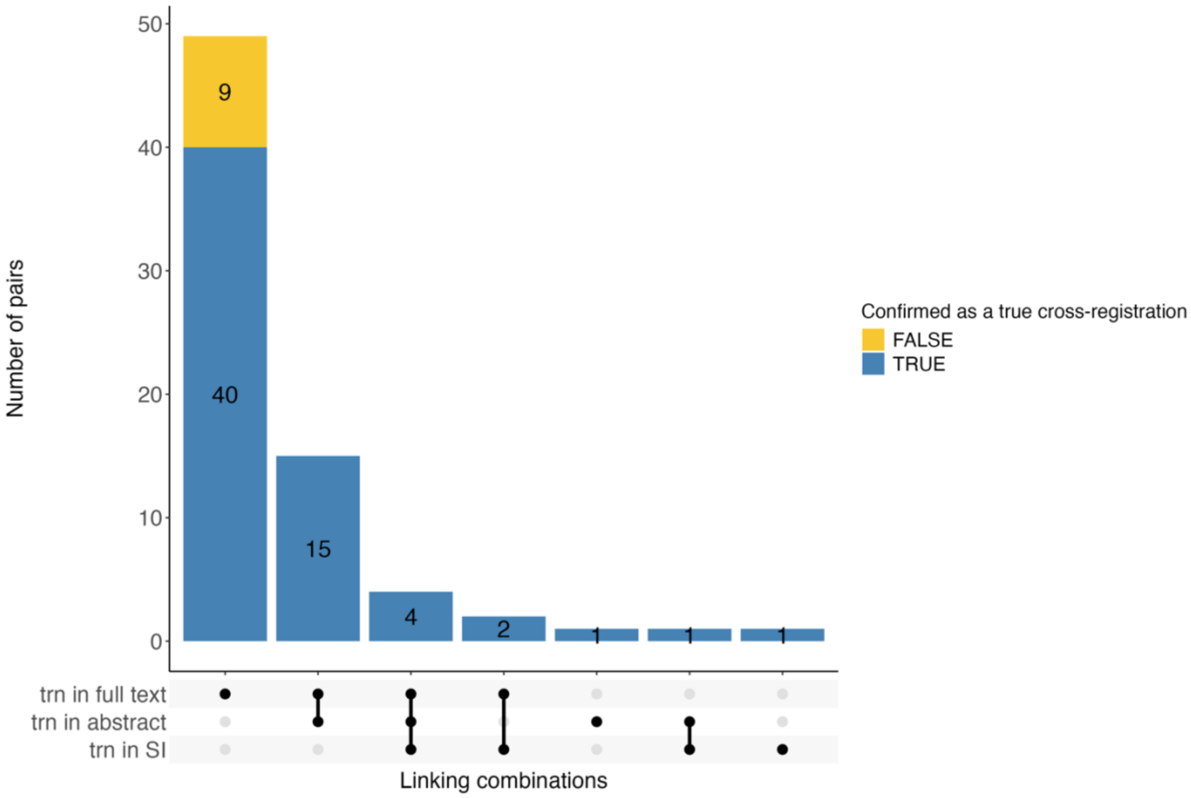


#### Potential cross-registrations involving duplicate TRNs

The 625 potential cross-registration pairs identified in this study involved 614 unique registrations from IntoValue (11 TRNs in IntoValue were each linked to 2 EUCTR TRNs) and 620 unique EUCTR registrations (5 EUCTR TRNs were each linked to more than one registration in DRKS or ClinicalTrials.gov). As outlined below, some of these registration pairs had already been included in the manual review. Following completion of the review, we systematically checked all registration pairs involving duplicate TRNs since they may be indicative of false cross-registrations:

- *11 ClinicalTrials.gov TRNs (IntoValue) were each linked to 2 distinct EUCTR TRNs, leading to a total of 22 registration pairs.* Of these, 11 were true positives and 11 were false positives. Thus, all 11 ClinicalTrials.gov TRNs could eventually be linked to a single cross-registration in EUCTR. Of the 22 registration pairs, 11 were included in the manual review performed in this study (2 were true cross-registrations, 9 were false cross-registrations).
- *5 EUCTR TRNs were each linked to more than one registration in DRKS or ClinicalTrials.gov (total of 10 registration pairs).* Of these, 5 registration pairs were included in the manual review performed in this study (3 were true cross-registrations, 2 were false cross-registrations). Upon further manual inspection after completion of the study, we determined

that 3 of the 5 EUCTR TRNs were each connected to a TRN in ClinicalTrials.gov and a TRN in DRKS, and appeared to be true cross-registrations across all 3 registries (EUCTR, ClinicalTrials.gov, and DRKS). The remaining 2 EUCTR TRNs were each connected to 2 distinct ClinicalTrials.gov TRNs. In both cases, one of the cross-registrations was true and the other false.

Thus, flagging potential cross-registrations involving duplicate TRNs can help to prioritize cases to include in a manual review, particularly in case it is not feasible to manually review all cases.

### Supplement S6: Discrepancy analyses

#### Prospective registration in ClinicalTrials.gov and DRKS

**Figure S6-1** displays the prospective registration status (on ClinicalTrials.gov or DRKS) of IntoValue trials with a confirmed EUCTR cross-registration (n = 228). Of the 213 confirmed EUCTR – ClinicalTrials.gov registrations, 62% (n = 133) were prospective in ClinicalTrials.gov. In turn, of the 15 confirmed EUCTR – DRKS cross-registrations, 73% (n = 11) were prospective in DRKS. Note that registration in the EUCTR is inherently prospective, as registration is linked to regulatory approval by national competent authorities. To assess whether a study was prospectively registered, we compared the date the study was first submitted to the registry with the start date given in the registry. We defined a trial to be prospectively registered if the trial was registered in the same or a previous month to the trial start date, as some registrations provide only a start month rather than an exact date.

**Figure S6-1.** Prospective registration status for IntoValue trials with a confirmed EUCTR cross-registration.

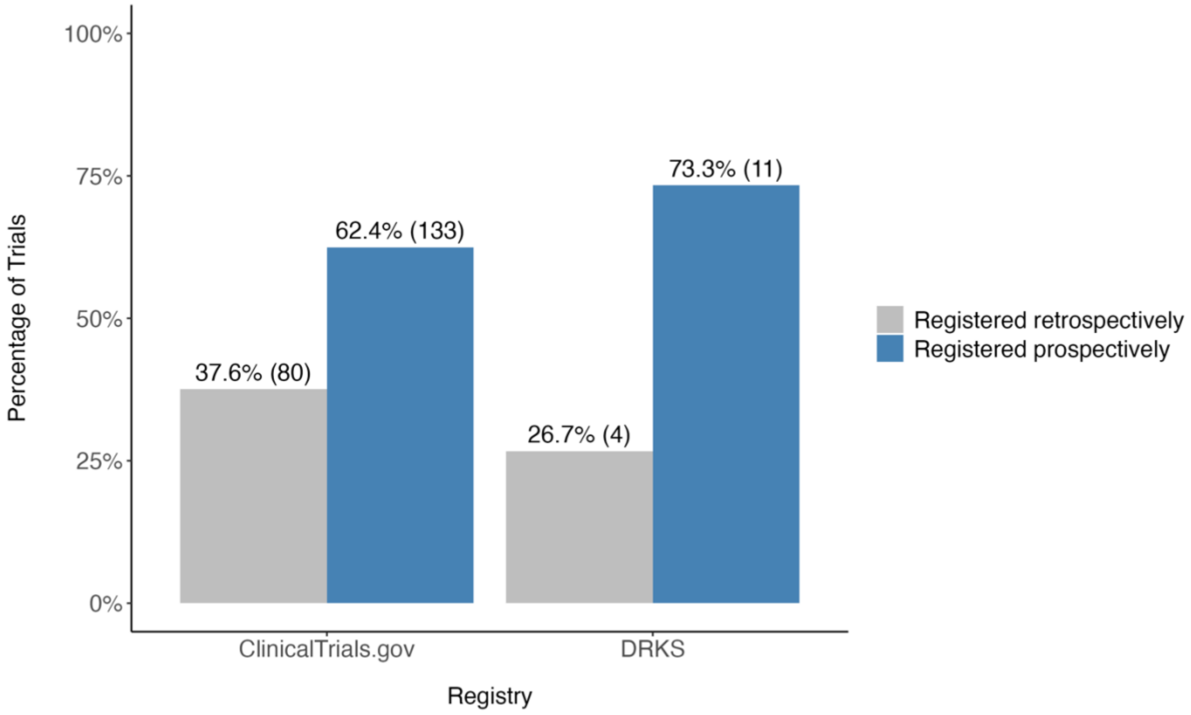

#### Recruitment status

**Table S6-1** displays the overall recruitment status for manually confirmed cross-registrations with an available recruitment status (n = 222). See **Table S2-1** for more information on how recruitment statuses in ClinicalTrials.gov, DRKS, and the EUCTR were mapped to an overall recruitment status in this study.

For confirmed cross-registrations between EUCTR – ClinicalTrials.gov (n = 207), 85% (n = 175) had the same overall recruitment status ("Completed"). The remaining 15% (n = 32) had a different overall recruitment status, with 8% (n = 17) "Completed" on EUCTR and "Other" on ClinicalTrials.gov, 5% (n = 11) "Ongoing" on EUCTR and "Completed" on ClinicalTrials.gov, and 2% (n = 4) "Ongoing" on EUCTR and "Other" on ClinicalTrials.gov. For confirmed cross-registrations between EUCTR – DRKS (n = 15), 93% (n = 14) had the same overall recruitment status ("Completed"), while the remaining case (n = 1) had an overall recruitment status "Ongoing" on EUCTR and "Completed" on DRKS.

**Table S6-1.** Overall recruitment status for manually confirmed cross-registrations with an available recruitment status (n = 222).

| | | **"Completed" on EUCTR** | **"Ongoing" on EUCTR** | **"Other" on EUCTR** |
|---|---|---|---|---|
| **"Completed" on other registry** | **(Subtotal)** | **189** | **12** | **0** |
| | ClinicalTrials.gov | 175 | 11 | 0 |
| | DRKS | 14 | 1 | 0 |
| **"Ongoing" on other registry** | **(Subtotal)** | **0** | **0** | **0** |
| | ClinicalTrials.gov | 0 | 0 | 0 |
| | DRKS | 0 | 0 | 0 |
| **"Other" on other registry** | **(Subtotal)** | **17** | **4** | **0** |
| | ClinicalTrials.gov | 17 | 4 | 0 |
| | DRKS | 0 | 0 | 0 |

#### Completion date: Sensitivity analysis

We performed a sensitivity analysis of the completion date, where completion dates in the EUCTR were taken solely from the protocol for Germany (i.e., EUCTR results pages were disregarded). Of the 195 confirmed EUCTR cross-registrations considered as "Completed" in both registries, 25 were excluded in this analysis due to a missing protocol for Germany in the EUCTR (n = 6) or because no completion date was available in the protocol in EUCTR (n = 19). Of the remaining 170 cross-registrations, 51% (n = 87; ClinicalTrials.gov n = 78; DRKS n = 9) had the same completion date across registrations. Focusing on discrepant cases, differences in completion date ranged from 1 – 42 months (median = 5, standard deviation = 7) (**Figure S6-2**). The completion date in EUCTR was earlier in 37% (n = 62) and later in 12% (n = 21) of cases.

**Figure S6-2.** Analysis of completion dates (month/year) across registrations of the same trial. Completion dates in EUCTR were only considered from the protocol for Germany.

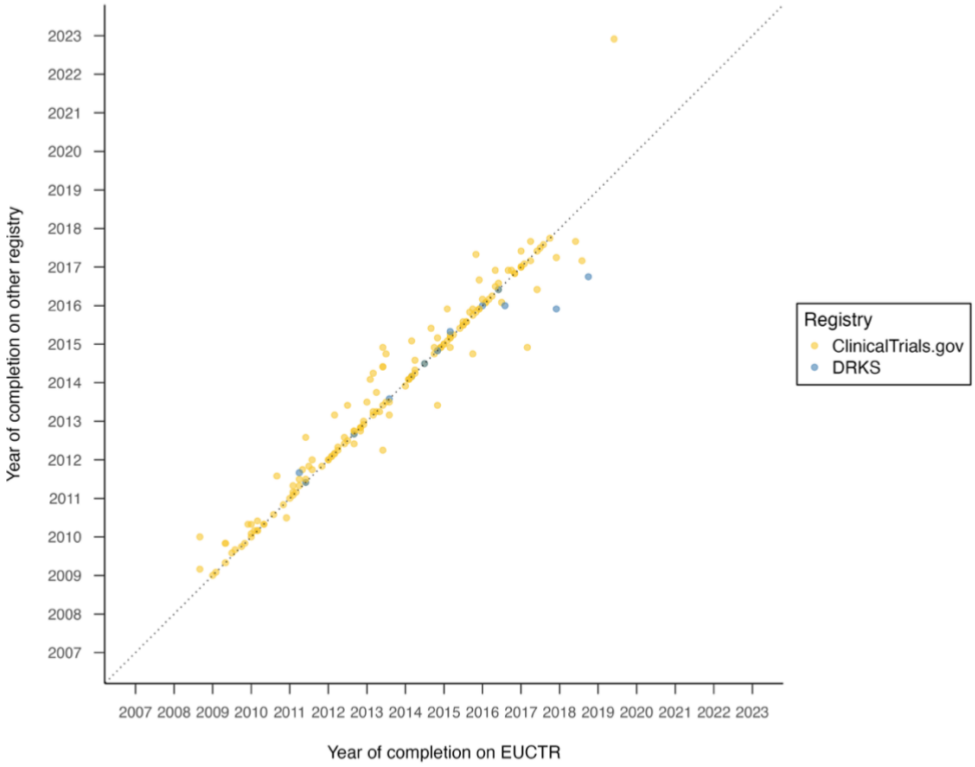

#### Summary results reporting: Sensitivity analysis

**Figure S6-3** builds on **Figure 5** (EUCTR – ClinicalTrials.gov) and displays reporting rates on the registry when broadening the definition of summary results to also include linked publications on ClinicalTrials.gov (in any field of the "Publications" section of the registration regardless of publication type). This further increased reporting to 89% (n = 161), with a little under half of cross-registrations (44%, n = 80) having posted on both registry entries. When assessing results reporting, we checked the type of report that was posted, but we did not perform a detailed review to confirm whether these reported on the results of the trial. See **Supplement S2** for an overview of reporting formats and their inclusion across analyses.

**Figure S6-3.** (Summary) results reporting for confirmed EUCTR – ClinicalTrials.gov cross-registrations across analyses.

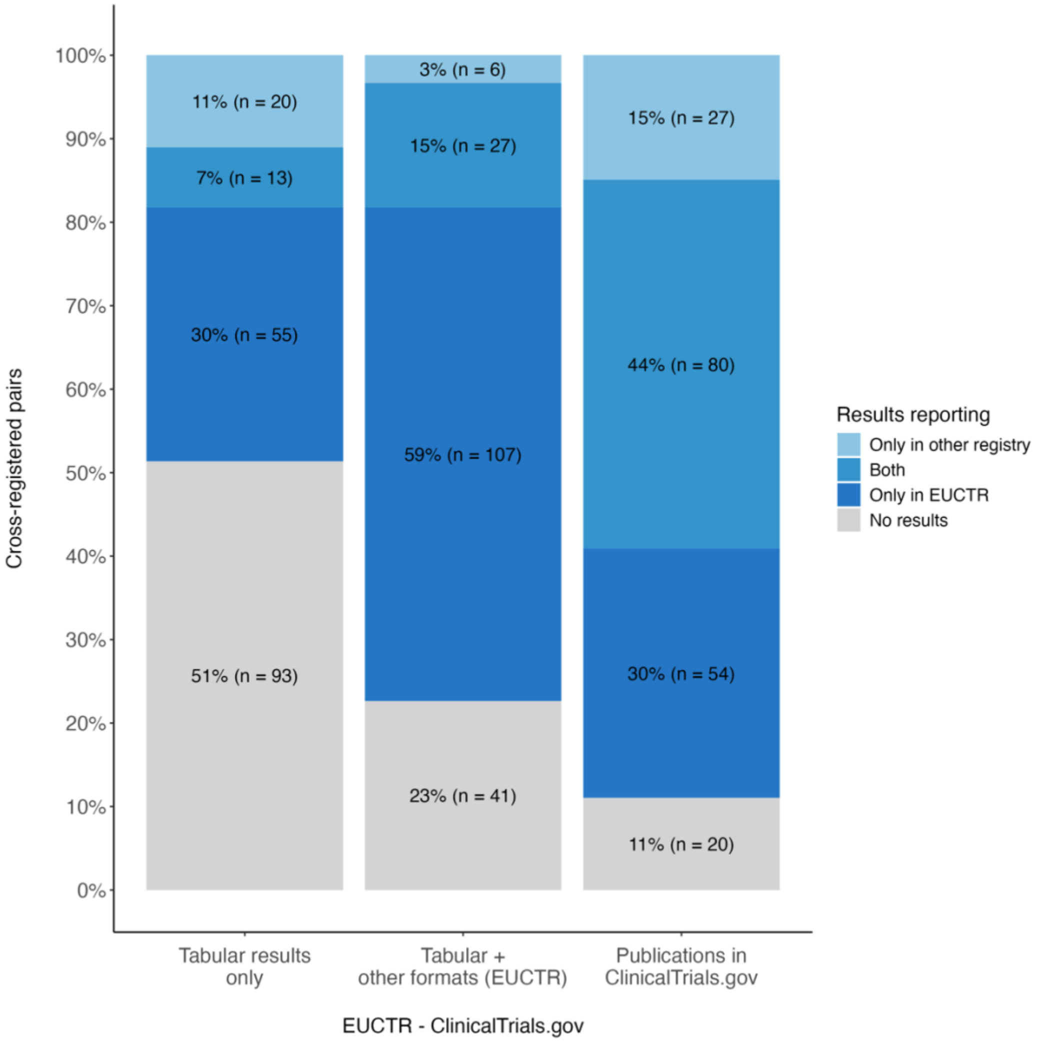
